# Targeted adaptive sampling enables clinical pharmacogenomics testing and genome-wide genotyping

**DOI:** 10.1101/2025.05.05.25326970

**Authors:** Pamela Gan Hui Peng, Yeo Han Lin, Muhammad Irfan Bin Hajis, Yusuf Maulana, Audrey Ng Qi Hui, Kevin Nathanael Ramanto, Marta Nisita Dewanggana, Astrid Irwanto, Levana Sani, Ling Goh Liuh, Mar Gonzalez-Porta

## Abstract

Pharmacogenomics (PGx) testing improves medication safety and efficacy by identifying genetic variants that affect drug response. However, current technologies often fail to resolve complex loci, detect structural variants, or phase alleles accurately. Here, we present an end-to-end PGx workflow based on Targeted Adaptive Sampling-Long Read Sequencing (TAS-LRS), integrating a streamlined laboratory protocol with a bioinformatics pipeline that includes a novel CYP2D6 caller. Using 1,000 ng of DNA and three-sample multiplexing on a single PromethION flow cell, the assay achieves consistent on-target (25x) and off-target (3x) coverage, enabling accurate, haplotype-resolved testing of 35 pharmacogenes alongside genome-wide genotyping from off-target reads. We further developed the workflow into a clinically ready service and validated its performance across 17 reference and clinical samples. The assay demonstrated high concordance for small variants (99.9%) and structural variants (>95%), with phased diplotypes and metabolizer phenotypes reaching 97.7% and 98.0% concordance, respectively. Improved calls were observed in 12 genes due to enhanced genotyping, phasing, or novel allele detection. In addition, off-target reads supported accurate genome-wide imputation, comparable to short-read sequencing and superior to microarrays. These results establish the feasibility of long-read sequencing for clinical PGx testing and position TAS-LRS as a scalable solution combining both targeted and genome-wide utility.

## 1 Introduction

Pharmacogenomics (PGx) is a branch of medicine focused on understanding how an individual’s genetic profile impacts their response to medications^1^. It examines the role of genetic variations in drug metabolism, effectiveness, and safety, aiding in understanding and predicting individual responses to specific drugs. PGx variants differ from those in other medical conditions in their high penetrance, widespread prevalence, and actionable potential. Although Adverse Drug Reactions (ADRs) are multifactorial in origin, a substantial subset, estimated at approximately one-third of serious cases, involve medications with known pharmacogenetic associations^2,3^. ADRs can manifest as allergic reactions or diminished drug efficacy, which can lead to negative outcomes or toxicity. For example, severe reactions such as Stevens-Johnson syndrome are associated to specific *HLA-B* alleles (*HLA-B*58:01* and *HLA-B*15:02*) in patients taking Allopurinol and Carbamazepine, respectively^4^. In terms of efficacy, certain *CYP2C19* alleles (*2 and *3) impair the metabolism of drugs like Clopidogrel, increasing the risk of cardiovascular events^5^. Similarly, individuals with extra copies of the *CYP2D6* gene metabolize codeine into morphine at an accelerated rate, enhancing pain relief but also heightening the risk of serious side effects, such as respiratory depression or, in extreme cases, overdose and death ^6^. PGx variants are also highly prevalent, with over 90% of the general population estimated to carry at least one variant that could significantly affect drug therapy ^1,7–9^. Lastly, and most importantly for clinical implementation, PGx variants are actionable. Consortia such as the Clinical Pharmacogenetics Implementation Consortium (CPIC) and the Dutch Pharmacogenetics Working Group (DPWG) have developed guidelines for over 100 gene-drug pairs widely available in the market^10,11^, which provide a robust and evidence-based framework for integrating PGx into clinical practice. These guidelines demonstrate that many ADRs can be mitigated through personalized approaches, such as dose adjustments or alternative therapies, highlighting the potential to improve effectiveness and safety without necessarily raising treatment costs. Indeed, several pilot studies have shown that pre-emptive PGx testing is feasible and can lead to reductions in emergency department visits, hospitalizations, and healthcare costs^12–15^. Among these, the PREPARE study - the largest prospective clinical trial to date conducted across seven European countries - reported a 30% decrease in ADRs in a diverse cohort of 6,944 patients^13^.

A range of technologies has been employed to characterize variation in PGx genes, including PCR, microarrays, and short-read sequencing^16^. More recently, long-read sequencing has emerged as a promising method for improving accuracy of genomic analysis^17^. Its ability to perform haplotype phasing without requiring parental data has the potential to aid in the interpretation of PGx variants ^18^, for example, by enabling the differentiation of genotypes such as *TPMT*3B/*3C* (where the *3B allele contains rs1800460 and *3C contains rs1142345) from *TPMT*1/*3A* (where *3A includes both rs1800460 and rs1142345). Long-read sequencing also offers higher resolution in complex genomic regions, such as structural variants and hybrid rearrangements, which are common in pharmacogenes like *CYP2D6*, *UGT1A1*, and *HLA* genes. For instance, the *CYP2D6* *36+*10 haplotype, a common variant in East Asians characterized by a hybrid tandem duplication, is difficult to detect with standard microarray chips^19^ and may also be missed by short-read sequencing approaches^20–23^. Lastly, long-read sequencing is not limited by prior knowledge of specific variants, making it capable of identifying rare and novel alleles - a common limitation of targeted methods such as PCR, microarrays, and sequencing panels. While historically more costly, the introduction of Targeted Adaptive Sampling-Long Read Sequencing (TAS-LRS) by Oxford Nanopore Technologies (ONT) has made long-read sequencing more accessible^24–26^. TAS-LRS leverages the real-time base-calling capabilities of nanopore sequencing to enrich for DNA fragments that map to a set of pre-defined genomic regions. The process begins by sequencing an initial stretch of each DNA molecule that enters the pore (approximately 400–800 bp). Basecalling and alignment are performed in real time to determine whether this short sequence matches the specified target regions. If a match is found, sequencing continues to generate a long read; if not, the fragment is ejected, allowing the next fragment to enter the pore. As a targeted method, TAS-LRS supports multiplexing, which reduces sequencing costs. Combined with the lower burden of instrument investment, this makes the technology more accessible to research and clinical laboratories. In addition, unlike conventional targeted assays that preselect DNA fragments during library preparation, TAS-LRS enriches target regions while also producing low-depth off-target data, enabling potential genome-wide analysis.

Here we introduce an end-to-end pharmacogenomics testing workflow based on TAS-LRS. We describe the experimental protocol and bioinformatics reporting pipeline, which integrates external tools with in-house algorithms, including a novel *CYP2D6* caller. We also outline the development of this workflow into a clinically ready service, presenting analytical and clinical validation results assessing the assay’s Limit of Detection (LOD), accuracy, precision, and specificity. In addition, we demonstrate the utility of off-target signals generated during adaptive sampling for genome-wide genotyping, highlighting the broader research potential of this approach. Altogether, our workflow enables, for the first time, the clinical implementation of PGx testing using long-read sequencing.

## 2 Results

### 2.1 TAS-LRS workflow and validation study design

The implementation of PGx into clinical practice requires an end-to-end workflow that ensures analytical validity, clinical utility, and operational feasibility. Here we present a clinical workflow for pre-emptive PGx testing based on TAS-LRS), designed to balance comprehensive variant detection with cost-efficiency and scalability. The workflow consists of four key phases: (i) pre-test consultation (optional), (ii) sample preparation and sequencing, (iii) bioinformatics analysis and reporting, and (iv) post-test consultation, with multiple Quality Control (QC) steps applied throughout (**Figure 1a** and **Methods**). The reporting workflow covers 35 pharmacogenes (**Supplementary Table 1**), including all Very Important Pharmacogenes (VIPs) from Pharmacogenomics Knowledgebase (PharmGKB) (N=34) along with *HLA-A*, which is routinely tested in our laboratory. Additionally, genome-wide imputation from off-target reads is used for broader genotyping, supporting downstream research applications. Beyond genetic variation, the workflow also captures base modification signals, including 5mC, 5hmC, 6mA, and 4mC, leveraging ONT’s native DNA sequencing capabilities. While not analyzed in this study, these epigenetic features offer potential insights into gene regulation and drug response for future research.

**Figure 1.**
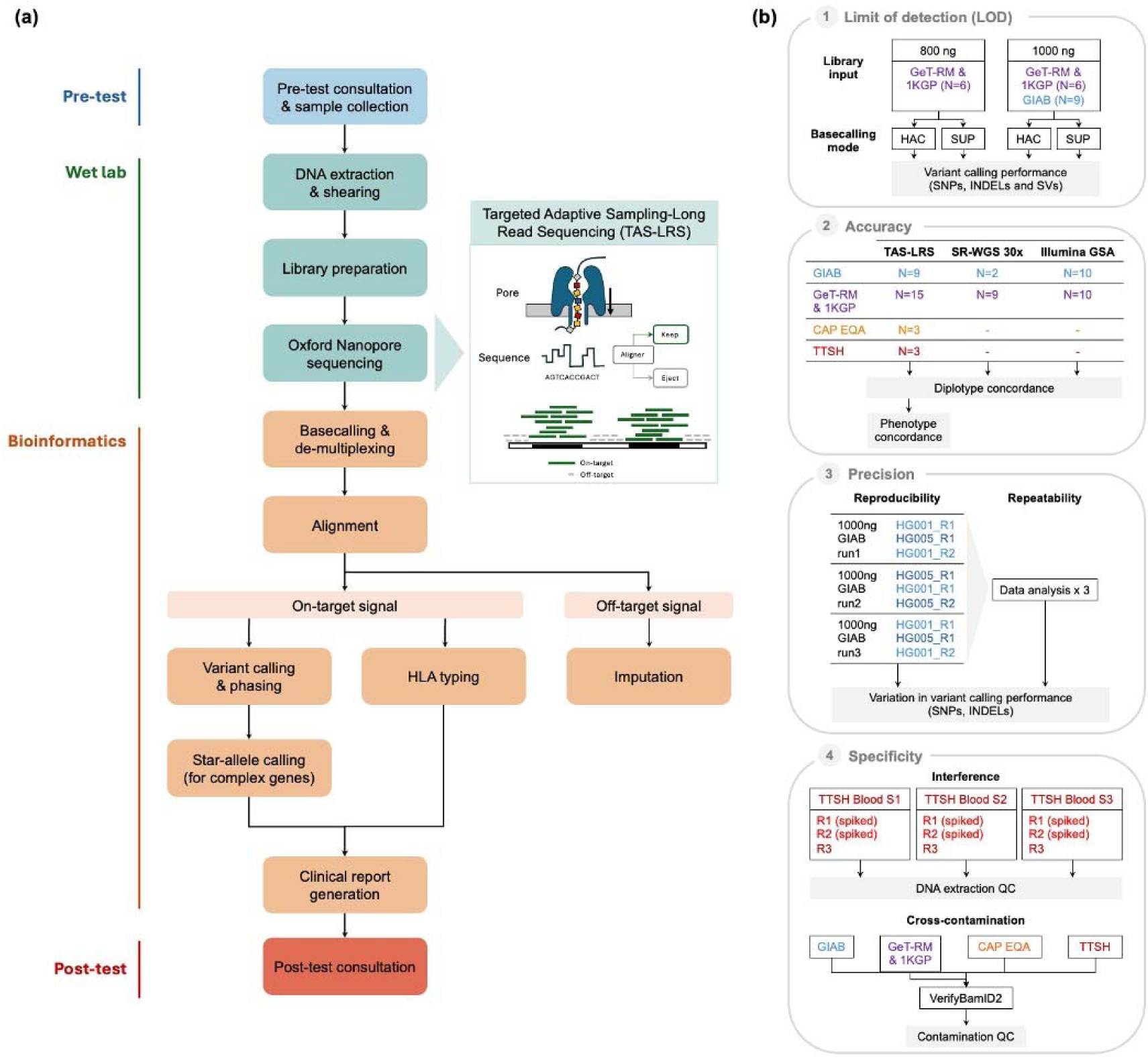
Overview of the TAS-LRS clinical workflow and validation framework. **(a)** Schematic of the end-to-end pharmacogenomics testing workflow based on Targeted Adaptive Sampling–Long Read Sequencing (TAS-LRS), comprising four main phases: optional pre-test consultation and sample collection; wet lab processing (DNA extraction, shearing, and Oxford Nanopore sequencing); bioinformatics analysis (including alignment, variant and star-allele calling, HLA typing, and genome-wide imputation); and post-test clinical consultation. Adaptive sampling enriches for 35 target pharmacogenes, including all PharmGKB Very Important Pharmacogenes (VIPs) and HLA-A, while generating low-depth off-target data for broader genome-wide applications. Blue, green, orange, and red indicate distinct workflow stages. **(b)** Validation framework for the TAS-LRS workflow, consisting of four core studies: (1) Limit of Detection (LOD), evaluating variant calling performance (SNVs, indels, and SVs) across different DNA input amounts (800 ng vs 1000 ng) and basecalling models (High-accuracy (HAC) vs Super-accuracy (SUP)); (2) Accuracy, assessed by diplotype and phenotype concordance across reference cell lines (GIAB, GeT-RM, 1KGP), external quality assessment (CAP EQA), and clinical samples from Tan Tock Seng Hospital (TTSH); (3) Precision, evaluated via reproducibility across sequencing runs and repeatability across independent bioinformatics analyses; and (4) Specificity, assessed through interference testing using triglyceride-spiked blood samples and cross-contamination testing using VerifyBamID2.

To evaluate the analytical performance of this workflow, we designed a validation framework based on recommendations from regulatory guidelines, including those from the Food and Drug Administration (FDA), College of American Pathologists/Clinical and Laboratory Standards Institute (CAP/CLSI), Clinical Laboratory Improvement Amendments (CLIA), and In Vitro Diagnostic Regulation (IVDR) ^27^. The framework consists of four studies assessing key performance metrics: LOD, accuracy against established truth sets, precision (reproducibility and repeatability) and specificity (interference and cross-contamination) (**Figure 1b**). The validation study included 17 unique samples across 10 sequencing runs (**Supplementary Table 2**), incorporating cell lines, CAP External Quality Assessment (EQA) samples, and real patient specimens to ensure clinically relevant testing. Cell lines were selected to represent a range of pharmacogenomic variants, including small variants (SNPs and indels) and structural variants (SVs). CAP EQA and patient samples were used to validate performance against orthogonal methods, including clinically validated workflows.

Across all validation runs, and when combining both the 1-hour Whole Genome Sequencing (WGS) and adaptive sampling stages, we obtained a mean sequencing yield of 37.09 Gbases per run, with a mean base quality score of 21.65. As expected, run yield positively correlated with the number of available starting pores on the flow cell (R² = 0.67, p = 0.0036 for total bases; R² = 0.66, p = 0.0043 for passed bases; Supplementary Figure 1). De-multiplexing efficiency for the adaptive sampling runs using reads with Q-score>10 was greater than 95%, resulting in an average of 10.38 Gbases of usable output per sample with our 3-plex strategy. On-target regions (N=326) achieved a mean coverage of 25.2x, while off-target regions had an average coverage of 3.0x, indicating a mean fold enrichment of 8.5 (**Supplementary Table 3** and Supplementary Figure 2). N50 values were as expected, with an average of 7,889 bp in on-target regions and 641 bp in off-target regions, confirming that adaptive sampling successfully enriched sequencing depth for the desired loci. Closer inspection of on-target coverage across the 35 pharmacogenes revealed a mean coverage of 21.9x (Supplementary Figure 3). Only two genes, *G6PD* and *SLC19A1*, consistently showed lower coverage. For *G6PD*, the reduced coverage can be attributed to its X-linked location, which results in sex-dependent coverage differences. Indeed, female samples, which are expected to have approximately double the copy number of males, achieved an average depth of 17x, while male samples reached only 10x. In the case of *SLC19A1*, the lower coverage may be due to a low-complexity region in the flanking sequence of the target region (Supplementary Figure 4), and could potentially be handled by further optimising padding in the target BED file.

### 2.2 Variant calling performance – Limit of detection (LOD) study

To evaluate the LOD of the TAS-LRS-based PGx workflow, we first assessed the accuracy of genotype calls using reference cell lines, focusing on raw genotype accuracy rather than diplotype interpretation. Given the potential impact of DNA input on long-read sequencing performance, we compared runs using 800 ng and 1000 ng of library input. Three runs were performed at 800 ng, while four runs were conducted at 1000 ng, including one sample (HG01190) sequenced under both conditions to enable direct comparison. Additionally, basecalling performance was evaluated using High-accuracy (HAC) and Super-accuracy (SUP) models.

For small variants, analysis included 1,142 variants (1,103 SNPs and 39 indels) across 31 PGx genes, excluding *CYP2D6*, *CYP2A6*, *HLA-A* and *HLA-B* due to their complex structural variation that impacts the accuracy of calls returned by Clair3 (see **Methods**). Across all conditions, and with SUP basecalling, callability averaged 99.49% and genotype concordance averaged 99.90%. No significant differences in performance were observed between DNA input conditions; however, basecalling with SUP instead of HAC resulted in improved performance, most notably for callability, which increased by 1.41% (**Table 1**).

For SVs, including deletions, duplications, and hybrid alleles, evaluations were performed at the individual event level, generating aggregate performance metrics across all samples (**Table 2**). Our analyses focused on *CYP2D6* and *CYP2A6*, as these were the only genes with SVs in our reference dataset of 17 unique samples. For duplications, performance remained consistent across DNA inputs and basecalling modes. For deletions, all expected events were also recovered, except for a missed *CYP2D6* *5 deletion in sample NA18861 when using 800 ng of input and HAC mode, which was likely due to the lower sequencing depth in this sample (18.22x), and which was successfully recovered when using SUP calling mode. Hybrid alleles presented a greater challenge, with performance differences observed between 800 ng and 1000 ng of DNA input. Specifically, an increased number of false negatives was detected at 1000 ng, consistently for HAC and SUP; however, further inspection suggested that sequencing depth, rather than DNA input alone, was the primary factor influencing this trend (27.4x mean depth in samples from 800 ng input runs vs. 24.2x in samples from 1000 ng input runs). Lastly, it is worth noting that the pipeline did not return any false positives, with specificity and precision consistently maintained at 100%.

These results highlight the strong influence of sequencing depth on SV detection, particularly for hybrid alleles. When performing a follow up analysis at a per-locus depth of 25x in *CYP2D6* and *CYP2A6*, genotype concordance reached 100%, indicating reliable detection across all SV types (**Supplementary Table 4**). The above findings also inform the choice of basecalling mode, with SUP demonstrating significant improvements in detecting both small variants and SVs. As a result, SUP was adopted as the default basecalling mode, and all subsequent validation studies were conducted using this approach.

### 2.3 PGx star allele diplotype and phenotype concordance – Accuracy study

While individual variant calling accuracy is critical for evaluating performance of genetic testing assays, pharmacogenomic workflows often require variant calls to be phased and interpreted into haplotypes, which are subsequently translated into metabolizer profiles. To address this, we expanded the performance assessment to evaluate the accuracy of diplotype and phenotype calling in pharmacogenes with complex alleles (i.e. star-alleles or similar; N=20). A significant challenge in this analysis lies in the extensive number of star-alleles associated with many pharmacogenes, exemplified by *CYP2D6*, which has over 100 known star-alleles catalogued in PharmGKB^28^. This diversity complicates the availability of reference materials, particularly for rare or population-specific haplotypes. To address these limitations, we selected samples representing a broad range of star-alleles, with particular focus on *CYP2D6*, one of the most highly variable pharmacogenes, and prioritized frequent haplotypes observed in Southeast Asian populations, such as *36+*10 hybrids (**Supplementary Table 2**). Cell lines from National Institute of Standards and Technology (NIST) and Coriell (N=11) were selected to include samples with truth data obtained either from Genetic Testing Reference Materials (GeT-RM), or orthogonal methods, such as 30x whole-genome short-read sequencing in the 1000 Genomes Project (1KGP). EQA samples were also included (N=3), with known expected results provided by previous CAP EQA outcomes. Finally, we also evaluated patient samples that had been previously tested using orthogonal methods (N=3), namely microarrays validated under a clinical workflow.

For each pharmacogene in each sample, diplotype calls generated by the TAS-LRS workflow were compared against truth data obtained from GeT-RM or orthogonal methods, including 30x whole-genome sequencing from 1KGP and clinically validated microarrays (see **Methods**). Diplotype concordance results were classified into four categories: *concordant*, where the call matched the truth set; *discordant*, where the call differed from the truth set; *improved*, where TAS-LRS calls were deemed correct upon manual review of both truth and test data; and *no-calls*, where the algorithm did not return a result due to quality control filters (see **Methods**). Overall, across 519 total diplotype calls evaluated, spanning 20 genes and 17 samples, we observed 446 concordant calls (85.93%), 2 discordant calls (0.39%), 61 improved calls (11.75%), and 10 no-calls (1.92%) (**Figure 2**). Aside from concordant calls, the next most frequent category was improved calls. We identified improvements in 12 out of 20 genes (60.00%), including *CYP2A6, CYP2B6, CYP2C9, CYP2D6, CYP4F2, DPYD, GSTP1, HLA-B, NAT2, NUDT15, SLCO1B1,* and *UGT1A1*.

**Figure 2.**
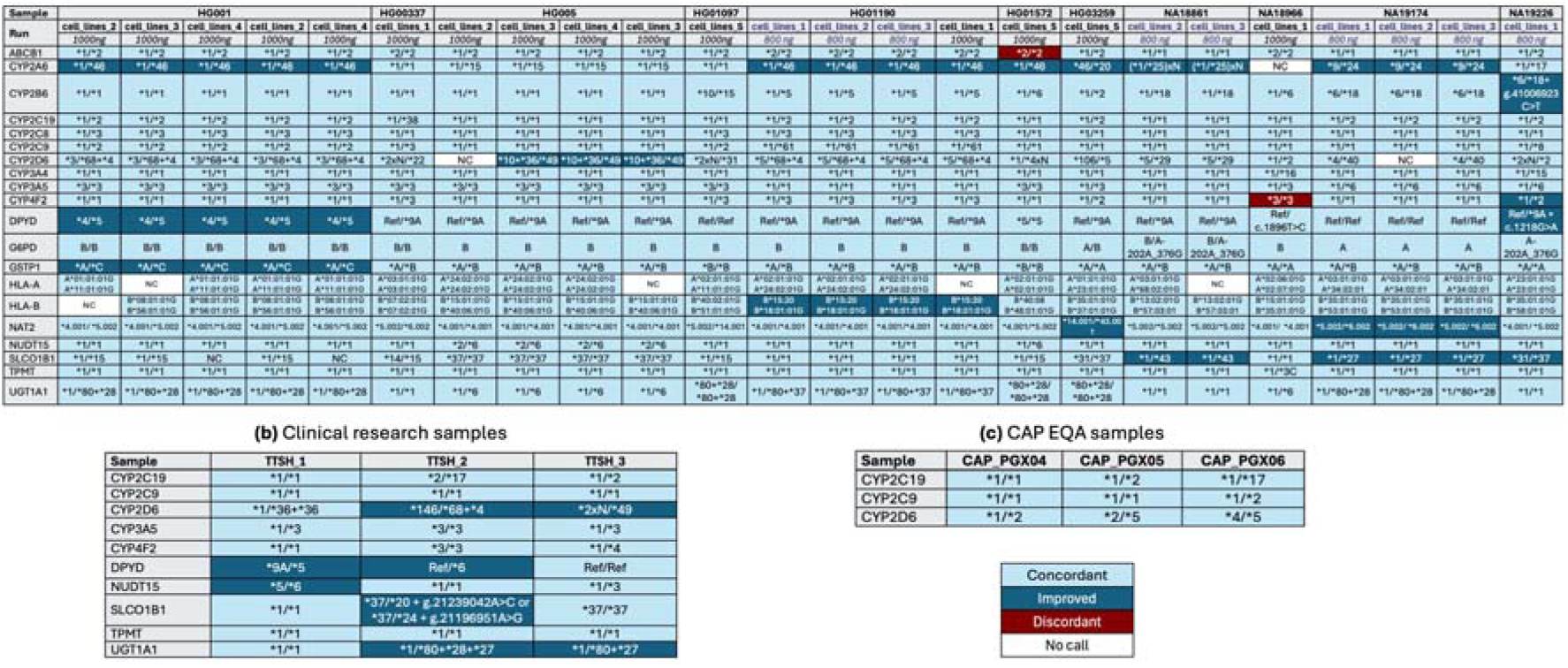
Diplotype concordance for pharmacogenes with complex alleles. Results of the TAS-LRS workflow for **(a)** reference cell lines (n=11), **(b)** clinical research samples (n=3) and **(c)** external quality assessment (EQA) samples (n=3) were compared against reference truth sets. GeT-RM calls were used as the truth set for HG001, HG01190, NA18861, NA18966, NA19174, and NA19226 unless otherwise specified. Diplotype calls for HG00337, HG005, HG01097, HG01572, and HG03259 were compared to PyPGx diplotypes from 30x short-read data. For all samples, *ABCB1, G6PD, NUDT15*, and *UGT1A1* calls were compared to PyPGx outputs, and *HLA-A* and *HLA-B* calls were evaluated against optitype v1.3.5. Calls matching the truth set were labelled as concordant, while mismatched calls were considered improved based on IGV read support or consensus across orthogonal methods.

These improvements were primarily driven by an expanded reportable range (N=32), more accurate genotyping (N=16), phasing (N=9) and SV detection (N=1), and identification of novel alleles due to enhanced phasing (N=3). Notably, improved calls remained consistent across replicates, further supporting their validity. For example, *NAT2* in sample NA19174 was ambiguously reported in the GeT-RM truth set as *4/*5 or *5/*6. This ambiguity persisted across short-read sequencing datasets analyzed with different tools, except for StellarPGx, which reported another set of potential star alleles (*16/*34). With TAS-LRS, the diplotype was fully resolved to *5.002/*6.002, with phasing supported by underlying read alignments in Integrative Genomics Viewer (IGV) (**Figure 3a**). Clinically, distinguishing *5/*6 (slow acetylator) from *4/*5 (intermediate acetylator) is critical, as *NAT2* encodes N-acetyltransferase 2, an enzyme involved in metabolizing drugs such as isoniazid, hydralazine, and sulfasalazine, and slow acetylators are at increased risk of drug-induced toxicities, including isoniazid-associated hepatotoxicity. Similarly, for *CYP2B6* in sample NA19926, both GeT-RM and orthogonal microarray and short-read datasets consistently returned a *18/*20 call, while PyPGx on short-read data suggested a possible novel allele that was not fully resolved. TAS-LRS phased the four variants involved in *18 and *20 into a novel configuration, assigning two variants to *6 in haplotype 1 and two variants to a novel allele in haplotype 2. These phasing patterns were clearly supported by IGV read alignments (**Figure 3b**). This reconfiguration has potential clinical implications, as *CYP2B6* alleles such as *6 and *18 are associated with reduced enzymatic activity, impacting the metabolism of drugs including efavirenz, bupropion, and methadone. In another similar case, TAS-LRS identified a novel *DPYD* haplotype in sample NA19226 that was not supported by short-read data or reflected in the GeT-RM truth set (Supplementary Figure 5a). While GeT-RM reported a *1/*9A diplotype based on the presence of c.85T>C (rs61622928), TAS-LRS revealed that c.1218G>A (rs1801265) co-occurred on the same haplotype. In this case, the ability to phase multiple functional variants on the same allele has important implications for fluoropyrimidine dosing, drugs that are used to treat colorectal and breast cancers and are primarily inactivated by *DPYD.* Both c.85T>C and c.1218G>A are missense variants associated with reduced *DPYD* enzyme activity in functional studies, and, when phased together on the same allele, their combined effect may further compromise enzyme function compared to either variant alone. As a final example, TAS-LRS resolved a complex *UGT1A1* diplotype in a clinical sample previously reported as *1/*28 based on microarray genotyping. Phased long-read data showed that the *28 allele co-occurred with *80 (rs887829) and *27 (rs35350960), yielding a corrected diplotype of *1/*80+*28+*27 (Supplementary Figure 5b). These additional variants, particularly *27, contribute to further reduced gene expression beyond the effect of *28 and misclassifying this diplotype as *1/*28 may therefore underestimate the risk of irinotecan-induced toxicity or hyperbilirubinemia.

**Figure 3.**
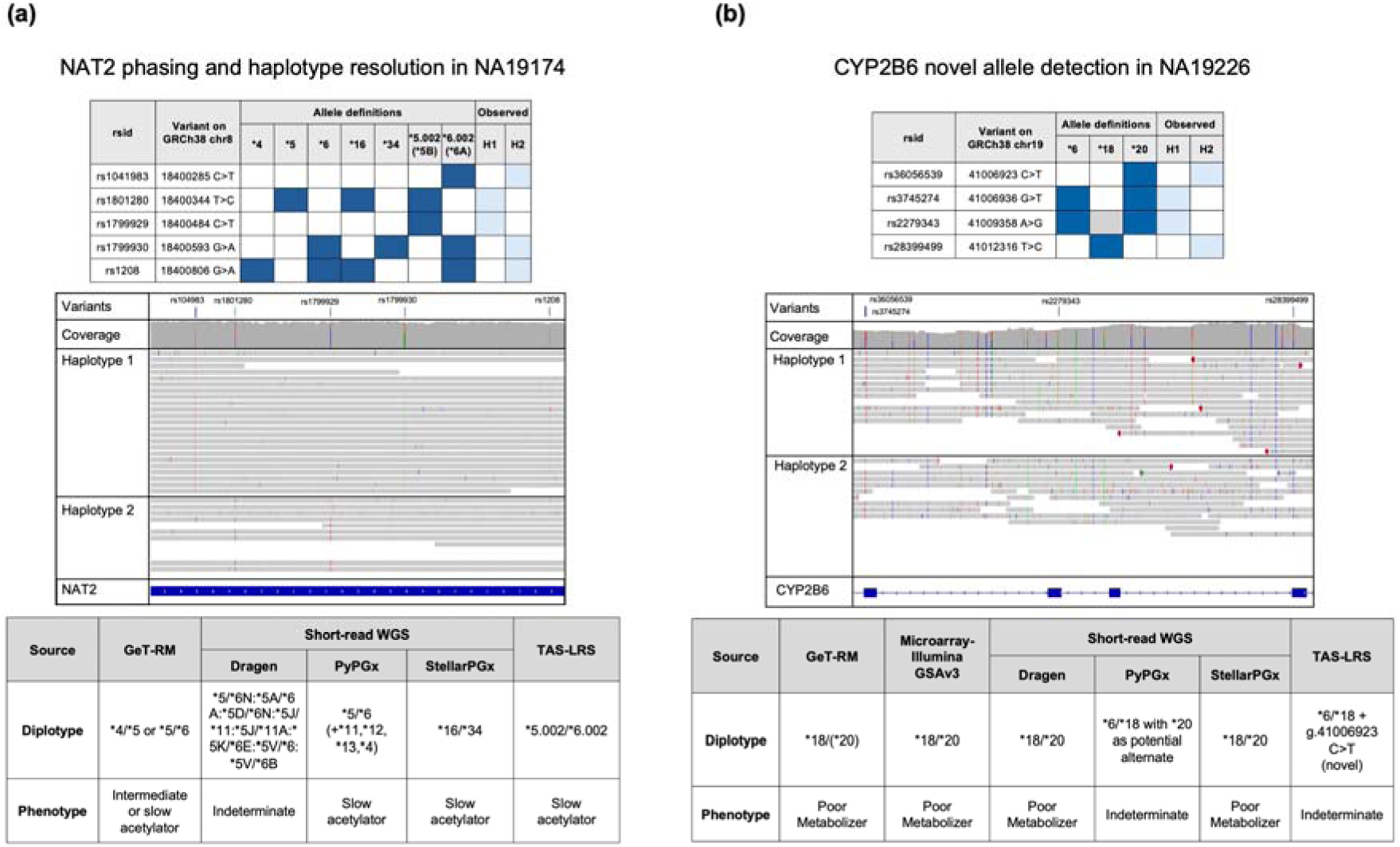
Examples of improved pharmacogene calls enabled by enhanced phasing, novel allele detection, and expanded haplotype coverage. **(a)** Phasing and haplotype resolution for *NAT2* in NA19174. **(b)** Improved phasing and detection of a novel allele in *CYP2B6* for NA19226. TAS-LRS resolved a partial call (*6/*18 + rs36056539) as the correct genotype.

In contrast to the improved calls, two discordant calls were observed in *ABCB1* and *CYP4F2*. For *ABCB1* in sample HG01572, Clair3 reported a TT genotype for the star-defining SNP rs2032582, which PyPGx interpreted as *2/*2, whereas short-read data supported a heterozygous alternate TC genotype, interpreted as *1/*2. IGV pileups showed evidence for the TC allele in TAS-LRS, but the C allele had insufficient support to be confidently called (**Figure 4a**). This suggests that increased sequencing depth at this locus would likely have resulted in the correct call. In *CYP4F2*, sample NA18966 was assigned a *3/*3 diplotype by TAS-LRS, whereas GeT-RM, microarray, and short-read data supported a *1/*3 call. Similarly to *ABCB1*, IGV analysis revealed that the incorrect call was due to low read depth, with very few reads spanning haplotype 1 (**Figure 4b**).

**Figure 4.**
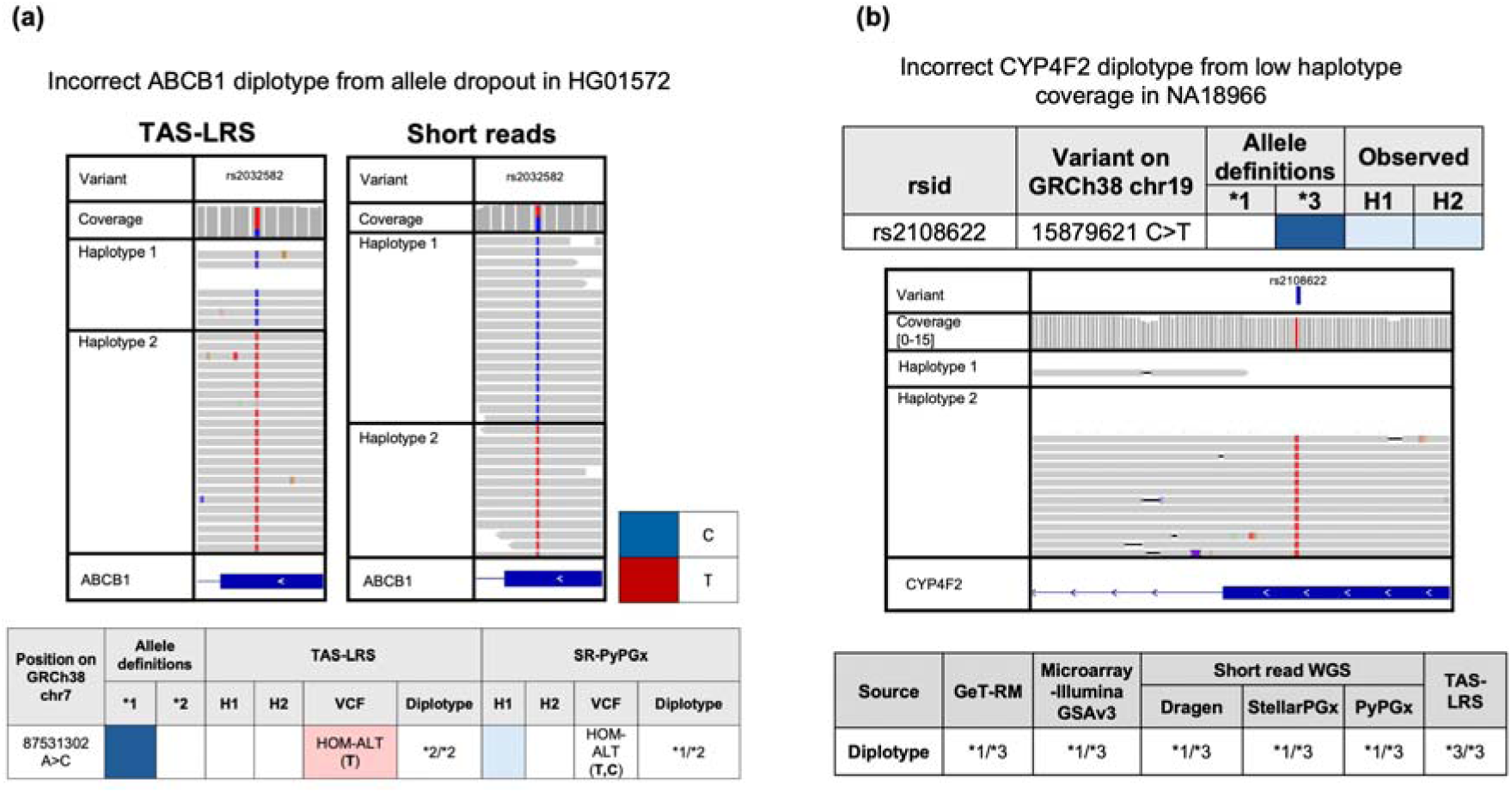
Examples of discordant calls caused by allele imbalance. **(a)** Discordant ABCB1 diplotype call in HG01572. rs2032582 was incorrectly called as homozygous alternate (TT) in long-read data, while short-read data supported a heterozygous genotype (TC). The C allele is present in the long reads, but its coverage is too low to meet the variant calling threshold. **(b)** Discordant CYP4F2 call in sample NA18966. TAS-LRS wrongly called the rs2108622 as homozygous alternate, whereas other platforms including GeT-RM, microarray (GSAv3), and short-read pipelines showed the diplotype as *1/*3. IGV plots show insufficient coverage of haplotype 1 for this specific sample.

Lastly, no-calls were observed in *CYP2A6*, *CYP2D6*, *HLA-A*, *HLA-B*, and *SLCO1B1*. For *CYP2A6*, a no-call arose from limitations in the current logic for combining a 3′ UTR conversion with a deletion; and algorithmic improvements are expected to resolve this. In *CYP2D6*, no calls were partly attributed to lower sequencing depth, where out of four HG005 replicates, the replicate with a no-call had the lowest sequencing depth (18x compared to a mean depth of 20.5x for the remaining samples). In addition, a no-call for NA19174 was observed despite having 32x coverage, due to a low-quality indel call at rs72549356, which did not match the expected variant. Note however that this indel was correctly detected in two other replicates at 37x and 26x. *HLA-A* and *HLA-B* no-calls were due to diplotype quality scores below the 0.95 threshold, also linked to insufficient depth. For *SLCO1B1*, no-calls were related to unresolved ambiguous diplotypes: in the two no-call replicates of HG001, the workflow identified two possible diplotypes, *1/*15 and *1/*46, which share two SNPs, rs2306283 and rs4149056, but differ in a third SNP, rs71581941, present only in *46; and due to insufficient evidence to detect the *46-defining SNP, it conservatively assigned a no-call.

In addition to the diplotype concordance analysis described above, in which results were compared against available truth data, we conducted a direct comparison between our TAS-LRS workflow and orthogonal testing platforms, including short-read 30x whole-genome sequencing (SR-WGS) analyzed with multiple bioinformatics workflows, and microarrays (Illumina GSAv3) (see **Methods**). Although limited to a subset of samples with available data (N=11), the analysis showed that TAS-LRS was able to achieve consistently high concordance rates (>95%), outperforming other platforms in 13 of 20 genes evaluated (**Supplementary Table 5**). Across platforms, genes with complex structural variation or multiple gene copies, such as *CYP2D6* and *CYP2A6*, and highly polymorphic genes like *UGT1A1* and *HLA-B*, posed particular challenges for both SR-WGS and microarrays. Microarrays, as expected, presented challenges with diplotype resolution for alleles requiring phasing or detection of SVs. While DRAGEN achieved the highest concordance among short-read workflows, it exhibited lower callability compared to TAS-LRS, whereas other pipelines, such as StellarPGx and PyPGx, showed high variability across genes. Importantly, short-read workflows often rely on population-based haplotype inference for diplotype assignment, introducing uncertainty when encountering novel or rare allele combinations. In contrast, TAS-LRS simplifies interpretation by directly delivering phased diplotypes from sequencing reads, minimizing reliance on statistical inference, and potentially reducing manual curation efforts in clinical laboratories.

Finally, since the PGx workflow ultimately generates metabolizer profiles for clinical reporting, we evaluated phenotype concordance across 13 genes with established phenotype interpretation tables. The classification system mirrored that used for diplotypes: *concordant* (matching the expected result based on either the reference or the improved diplotype call), *discordant* (not matching the expected result), or *indeterminate*. As expected, and consistent with diplotype concordance results, the majority of phenotypes were concordant (341/348, 97.99%), with only seven cases (2.01%) classified as indeterminate due to a no-call or insufficient copy number information, and no discordant phenotypes observed (Supplementary Figure 5). We subsequently focused on samples previously identified to have improved diplotype calls (**Figure 2**), since these are the ones more likely to lead to conflicting phenotype interpretations. Among the 52 improved diplotypes across the 13 genes evaluated, the majority (n=36, 69.23%) had no impact on phenotype interpretation. However, in 10 cases (19.23%), phenotype assignment was not possible due to the presence of novel alleles with uncharacterized function. This highlights a key trade-off of higher-resolution sequencing technologies: while they improve variant detection, they also increase the likelihood of identifying previously unreported alleles that require functional characterization before clinical recommendations can be made. These findings emphasize the need for ongoing curation efforts, and in the future, approaches such as *in silico* functional prediction or targeted functional assays could aid in assigning provisional phenotype classifications and further enhance clinical utility. Finally, six cases (11.54%) resulted in improved phenotype classification. An example is shown in Supplementary Figure 7, where TAS-LRS resolves a previously unphased copy number change in CYP2D6 for a clinical sample tested with microarrays, allowing for an unambiguous phenotypic interpretation. Without accurate resolution, this genotype could have led to inappropriate drug therapy recommendations and potential adverse outcomes.

### 2.4 Precision and specificity studies

To evaluate the precision of our assay, we performed reproducibility and repeatability studies using a subset of validation runs. Reproducibility was assessed by analyzing both inter- and intra-run consistency using GIAB replicates sequenced in three different runs (**Supplementary Table 3**). Across all performance metrics evaluated, inter-run and intra-run coefficients of variation remained below 5% (**Supplementary Table 6**), demonstrating high reproducibility. Repeatability was tested by running the analysis pipeline three times on the same input data from the same GIAB runs and starting from basecalling. Results were fully consistent across all repetitions, confirming the absence of stochastic variability in the analysis workflow.

Similarly, specificity was examined through interference and cross-contamination studies. To assess potential interference, three distinct blood samples were spiked with triglycerides at a concentration three times the optimal threshold based on the Singapore Ministry of Health 2016 Hyperlipidemia Guidelines (>450 mg/dL), simulating patients with severely elevated triglyceride levels (see **Methods**). Each sample was processed in triplicate, with two spiked replicates and one non-spiked control. While all non-spiked samples met the minimum DNA concentration requirements for downstream processing (min. 100 ng/ul for 3-plex sequencing), none of the spiked samples did (**Supplementary Table 7**). These results indicate that the test is not recommended for patients with significantly elevated triglyceride levels, as it may result in higher failure rates. This limitation should be clearly noted in clinical protocols and pre-test consultations, where severely elevated triglyceride levels could be considered an exclusion criterion or flagged for alternative sampling methods. No workaround, such as dilution or alternative extraction kits, was attempted in this study; however, future work may explore such options. Lastly, cross-contamination was assessed using VerifyBamID2^29^ to detect human DNA contamination in each sample. No signs of contamination were observed, with FREEMIX values consistently ≤3%, confirming high specificity (**Supplementary Table 8)**.

### 2.5 Off-Target signal analysis and genome-wide genotyping

Lastly, we investigated the potential use of off-target signal, generated from reads not selected during the adaptive sampling process (**Figure 1a**), to obtain genome-wide genotype calls. This capability is unique to the TAS-LRS workflow, as conventional targeted assays typically limit coverage to predefined regions, thus limiting genome-wide analysis. In our dataset, the mean off-target sequencing depth across all runs was 3.0x (**Supplementary Table 3**). Previous studies using both short-read and long-read sequencing have demonstrated that genome-wide genotyping from such low-coverage data is feasible through imputation techniques^30,31^. To assess this, we used the 1KGP reference panel to impute genotype calls in sample HG005 (Han Chinese), which is not part of the panel, to avoid bias. Imputation was performed using GLIMPSE, with results compared to those obtained from short-read sequencing downsampled to an equivalent depth and from a microarray dataset (see **Methods**).

Our results show that imputation based on the off-target TAS-LRS signal (mean coverage of 2.0x for the selected HG005 replicates) achieved high accuracy, closely matching that of short-read datasets and outperforming microarrays, particularly for rare variants with a minor allele frequency (MAF) <1% (**Figure 5**). This outcome highlights the unbiased nature of sequencing-based methods, which randomly sample DNA fragments across the genome, in contrast to microarrays that rely on predefined marker panels often biased toward common variants. Furthermore, the comparable imputation accuracy between short-read and long-read datasets demonstrates that, despite the lower base quality of ONT reads, TAS-LRS can generate equally reliable variant calls from imputation.

**Figure 5.**
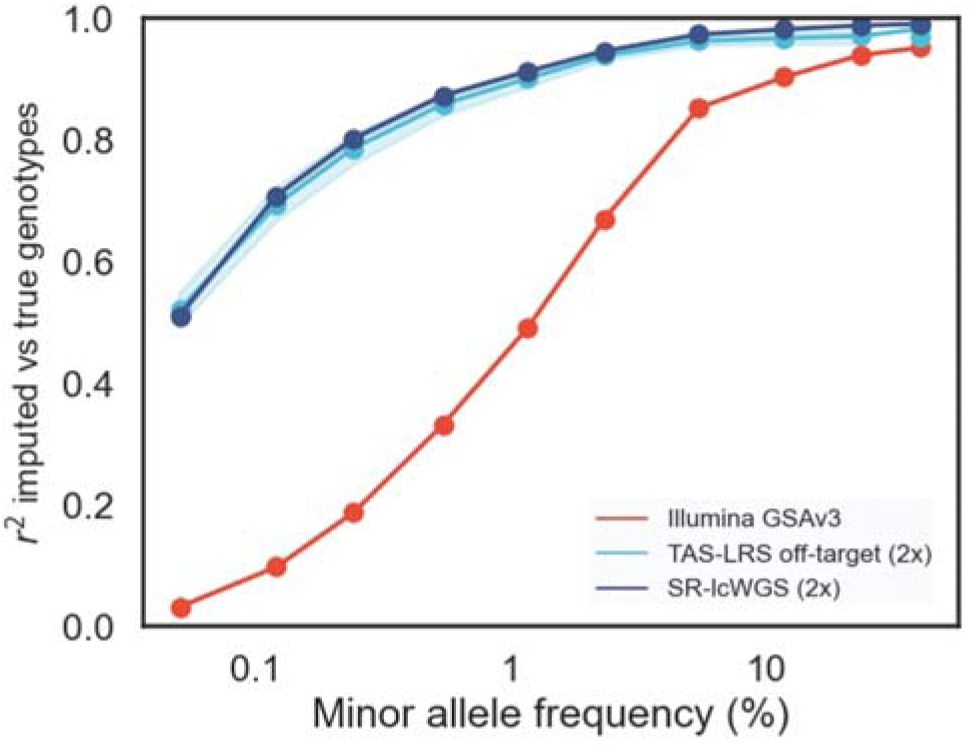
Genome-wide imputation accuracy across technologies. Imputation was performed using GLIMPSE2 (for TAS-LRS and short-read WGS) and Minimac4 (for microarray data), with the 1KGP reference panel which excludes the test samples (HG002 and HG005 from GIAB). Line plots show imputation accuracy (R²) across allele frequency bins. Rare variants are defined as those with minor allele frequency <1%. Shaded areas indicate 95% confidence intervals across replicates.

## 3 Discussion

Here, we introduce and validate an end-to-end pharmacogenomics (PGx) workflow based on targeted adaptive sampling long-read sequencing (TAS-LRS), developed to support clinical implementation in a CAP-accredited setting. The workflow combines an optimized laboratory protocol with a custom bioinformatics pipeline capable of resolving small variants, structural variants, and complex star alleles across 35 clinically relevant pharmacogenes, while also enabling genome-wide genotyping from off-target reads. Long-read sequencing technologies have gained increasing traction in recent years (i.e. they were named Method of the Year in 2023^17^) due to their ability to resolve complex genomic loci previously inaccessible with short-read sequencing. This capability is particularly relevant in PGx, where many pharmacogenes are highly polymorphic and reside in structurally challenging regions of the genome, often involving segmental duplications, tandem repeats, and pseudogenes^32^. A prominent example is *CYP2D6*, which encodes the cytochrome P450 2D6 enzyme responsible for metabolizing approximately 20% of clinically prescribed medications. To date, the *CYP2D6* locus includes over 170 catalogued star alleles, encompassing gene deletions, duplications, and hybrid rearrangements involving the closely related pseudogenes *CYP2D7* and *CYP2D8*^28^, with certain tandem hybrids, such as *36+*10, observed in up to 30% of East Asian populations^33–35^. Several prior studies have demonstrated the feasibility of using long-read platforms such as Oxford Nanopore and PacBio to resolve complex PGx loci, including early proof-of-concept work by Liau et al.^36,37^, Charnaud et al.^38^ and Turner et al.^22^ for *CYP2D6*, and by Graansma et al.^39^ for *CYP2C19*. Most recently, Deserranno et al.^25^ showcased the potential of nanopore adaptive sampling for PGx profiling across more than 1,000 genes. However, such efforts have been limited in sample size, gene coverage, or lack of formal validation frameworks. To our knowledge, ours is the first study to present a comprehensive analytical and clinical validation of a long-read-based PGx workflow, establishing key performance metrics (including LOD, accuracy, precision and specificity) to support its routine use in clinical diagnostics.

By analysing data from 10 sequencing runs comprising 17 unique reference and clinical samples, we demonstrated that our optimized adaptive sampling protocol yields consistent coverage, with a mean on-target depth of 25.2x and off-target depth of 3.0x, corresponding to an average enrichment of 8.5x, in line with expectations for TAS-LRS. We evaluated the impact of DNA input and basecalling mode on variant calling performance and observed that the SUP model consistently outperformed HAC for both small variants and structural variants in the target pharmacogenes. Notably, structural variant detection was more sensitive to sequencing depth, with 25x sufficient to recover all expected events in our dataset. However, further calibration is needed before establishing this as a universal threshold for clinical applications. To assess the clinical relevance of our workflow, we evaluated star allele and phenotype concordance across the 35 target pharmacogenes. Of 520 total diplotype calls, 507 (97.5%) were either concordant with the expected result (n = 446) or classified as improved upon manual curation (n = 61). Only two discordant calls (0.4%) and ten no-calls (1.9%) were observed, all attributable to insufficient coverage, which can be addressed through further protocol optimizations, such as improved DNA shearing, increased library loading and algorithmic refinements. To address loci prone to coverage variability (e.g., *G6PD*, *SLC19A1*), we recommend repeat sequencing or supplementary testing in cases where critical alleles cannot be confidently resolved. In addition, in a clinical setting, no-calls and ambiguous results should undergo review by a clinical scientist, with additional testing performed as necessary to ensure the reliability of reported genotypes.

As shown in previous long-read studies^38,40^, our findings underscore that accurate haplotype reconstruction, and not just variant detection, is essential for clinical PGx, particularly in genes with complex allelic architecture. For example, the *NAT2* diplotype in sample NA19174 was ambiguously reported as *4/*5 or *5/*6 in short-read datasets and reference truth sets. TAS-LRS resolved this to *5.002/*6.002, with phasing clearly supported by IGV alignments. Similarly, in *CYP2B6*, we observed an alternate phasing of four variants initially assigned to *18/*20, revealing a novel allele combination with well-supported haplotype structure. However, a key challenge remains in interpreting novel diplotypes for clinical actionability. While most improved calls (69.2%) maintained consistent phenotype interpretations, 10 of the 52 evaluated (19.2%) could not be mapped to known metabolizer categories due to the presence of novel or rare alleles. This limitation has also been noted in previous long-read PGx studies^39^, where increased resolution reveals variants not yet accommodated by current clinical guidelines. In clinical practice, novel or uncharacterized diplotypes would be reported with annotation of uncertain significance, with parallel efforts to expand reference databases and generate functional evidence to support future clinical interpretation. Conversely, in six cases (11.5%), improved genotyping enabled more definitive phenotype classification. For instance, in a clinical sample previously tested with microarrays, TAS-LRS resolved a copy number event in *CYP2D6* that had previously been unphased, enabling unambiguous assignment of metabolizer status.

While most of our validation focused on benchmarking against established truth sets and clinical workflows, we also compared TAS-LRS to orthogonal testing platforms where matched data were available. Current PGx genotyping technologies largely rely on microarrays or short-read sequencing, and therefore we included both in our evaluation. We observed that TAS-LRS achieved high concordance rates (>95%) and outperformed other platforms in 13 of the 20 genes assessed. As expected, microarrays showed the greatest limitations in resolving PGx diplotypes, consistent with prior reports^19^. It is important to note that we used the Illumina Global Screening Array v3 (GSAv3) chip, which is not specifically optimized for pharmacogenomic applications, and that newer arrays and PGx-focused designs may offer better performance. Nonetheless, microarrays inherently depend on prior knowledge of which variants to interrogate and are limited in their ability to discover novel alleles. Conversely, short-read sequencing performed better than microarrays but showed substantial variability across analysis pipelines. Among the evaluated algorithms, DRAGEN achieved the highest concordance, albeit with lower callability, a trade-off that could potentially be improved through further algorithmic refinement. Other notable short-read genotyping tools, such as Aldy 4^41^ and BCyrius^42^, were not included in this evaluation but warrant future benchmarking. However, phasing is likely to remain a limitation for short-read methods, as they rely on computational approaches based on population-level haplotypes, which are less reliable for individual-level predictions, particularly in admixed or ancestrally diverse populations. In contrast, long-read sequencing enables direct phasing of complex diplotypes and detection of novel allele configurations at the individual level. For example, we resolved previously uncharacterized phasing patterns in *NAT2*, *CYP2B6* and *DPYD* that were not captured by short-read approaches. Overall, our findings align with those reported by Barthélémy et al. (2023)^23^, who conducted a comparison of PacBio HiFi and Illumina short-read sequencing across a panel of pharmacogenes, and demonstrated that long-read sequencing not only improved haplotype resolution but also enabled accurate detection of structural variants, such as complex *CYP2D6* rearrangements and *UGT1A1* promoter repeats, that were miscalled or missed entirely by short-read methods. Furthermore, it is worth noting that our study was based on a relatively well-characterized sample set, with only three clinical specimens included. The frequency of novel or population-specific alleles is likely to be higher in routine clinical cohorts. For instance, in a previous study of approximately 1,800 Singaporean individuals of Chinese, Malay, and Indian ancestry, we found that ∼1% carried potentially novel *CYP2D6* haplotypes based on short-read whole-genome sequencing^35^. As long-read sequencing is applied to more diverse populations, we anticipate the discovery of additional novel variants that may have clinical relevance in pharmacogenomic decision-making.

In addition to enabling high-resolution PGx genotyping, TAS-LRS generates off-target reads that can be leveraged for genome-wide imputation. Imputation from low-pass sequencing is well established in the context of short-read data, and here we apply similar approaches to off-target signal from long-read sequencing^30^. Our results show that imputation accuracy from TAS-LRS off-target reads is comparable to that of short-read low-pass sequencing and exceeds that of microarrays, particularly for rare variants with a minor allele frequency (MAF) below 1%. This expands the utility of the assay beyond targeted PGx testing, enabling additional applications such as Polygenic Risk Score (PRS) development. Supporting this potential, Nakamura et al. (2024)^31^ demonstrated that PRSs derived from off-target long-read data in 33 hereditary cancer patients achieved accuracy comparable to those calculated from short-read sequencing. Altogether, these findings highlight the added value of adaptive sampling over conventional targeted sequencing methods, which typically rely on fixed panels enriched through PCR amplification or hybridization-based capture workflows that require custom library preparation. In contrast, adaptive sampling allows flexible updates to target content via simple BED file modifications, avoiding the need to redesign or reorder panels. This reduces wet-lab complexity, hands-on time, and consumable costs, and positions TAS-LRS as a versatile platform adaptable to a range of clinical contexts, including primary care, oncology, cardiovascular disease, and psychiatry.

In conclusion, our work demonstrates a robust clinical implementation of PGx testing using adaptive-sampling-based long-read sequencing. TAS-LRS combines accurate detection of small and structural variants, phasing without the need for parental data, and the ability to identify novel alleles, which are critical capabilities for PGx implementation given the complexity of pharmacogenes and the importance of haplotype resolution in predicting drug response. Beyond analytical performance, the simplicity and flexibility of the TAS-LRS protocol make it well suited for mainstream clinical adoption. Unlike fixed-panel assays, TAS-LRS allows rapid reconfiguration of target content without extensive redesign of experimental protocols, while simultaneously generating genome-wide data suitable for imputation. From DNA extraction to final report generation, the TAS-LRS workflow can typically be completed within 3 to 4 days, with faster turnaround possible for smaller batches or with higher parallelization. This positions TAS-LRS as a flexible platform that bridges targeted testing and broader genomic analysis, making it particularly compelling for national screening programs where both clinical relevance and research utility are priorities. In this context, and looking ahead, a unified assay integrating pharmacogenomics, polygenic risk, and monogenic variant analysis with potential for regulatory annotation through methylation profiling could enable comprehensive risk stratification from a single sample. Moreover, while long-read sequencing has historically been limited by cost, adaptive sampling enables multiplexing to reduce per-sample expenses. In this study, we demonstrated three-sample multiplexing, with four-sample runs already feasible and robust using current protocols. With further scaling and improved throughput, the cost of TAS-LRS could approach that of short-read whole-genome sequencing and may undercut combined short-read PGx panel and array workflows, especially as efforts to support five or more samples per run continue. Ultimately, the choice between long-read and short-read technologies will depend on context, scale, and clinical need; however, our findings demonstrate that long-read sequencing has matured beyond a research tool and now represents a viable option for clinical pharmacogenomics at scale.

## 4 Materials and Methods

### 4.1 End-to-end workflow for pre-emptive PGx testing

Our pre-emptive PGx workflow follows four main steps: (i) pre-test consultation (optional), (ii) sample preparation and sequencing, (iii) bioinformatics analysis and reporting, and (iv) post-test consultation (**Figure 1**). As this study focuses on generating evidence for implementation in a clinical laboratory, steps 1 and 4 were not performed but details are included below for completeness.

#### 4.1.1 Pre-test consultation (optional)

The patient eligibility assessment is a critical step in the TAS-LRS workflow to ensure appropriate test use. Eligibility criteria comprise excluding patients with conditions that may interfere with DNA integrity, such as recent blood transfusions or stem cell transplants, as well as those with impaired liver function, including patients with autoimmune hepatitis or a history of liver transplantation. For eligible patients, pre-test consultation is optional and offered based on institutional policy or clinician discretion. When provided, it includes counselling on the scope of PGx testing, potential actionable findings, and implications for medication management. Informed consent is obtained prior to sample collection.

#### 4.2.2 Sample preparation and sequencing

Sample preparation and sequencing follow the standard ONT laboratory workflow, optimized for multiplexing, as summarized in Supplementary Figure 8. The detailed protocol is described below.

##### Whole blood extraction

Whole blood samples were collected in K2EDTA tubes (VACUETTE) with a minimum volume of 3 mL. DNA was extracted from 300 µL of whole blood using the Monarch Genomic DNA Purification Kit (New England Biolabs), following the manufacturer’s protocol for non-nucleated mammalian whole blood with several modifications to accommodate the increased requirements for input volume for long-read sequencing (Supplementary Figure 9): (i) the lysis mastermix was scaled up threefold, and the lysis incubation time was extended to 25 minutes; (ii) after lysis, samples were split into two parts, with 1.5x the volume of binding buffer added per part; (iii) following binding, DNA from both fractions was concentrated into a single spin column and processed through the standard washing steps; and (iv) a double elution with preheated elution buffer was performed to maximize recovery. DNA yield and purity were assessed using the Nanodrop 2000, with a mean yield of 46.53 ng/µL across all samples.

To meet the yield requirement for TAS-LRS 3-plex (>100 ng/µL), the extracted genomic DNA (gDNA) was further concentrated using Solid-Phase Reversible Immobilization (SPRI) beads from MGI Tech Co., Ltd. (Supplementary Figure 10). Note that a lower DNA input is feasible at alternative plexities, and this is currently under investigation. A 2x volume of SPRI beads was added, and DNA was eluted in 30 µL of elution buffer. Final DNA quantification was performed using the 1x double-stranded DNA (dsDNA) High Sensitivity Qubit Assay (Thermo Fisher Scientific), yielding a mean concentration of 108.3 ng/µL per sample.

##### Genomic DNA shearing

Genomic DNA from cell lines and extracted blood samples was mechanically sheared to an average fragment length of 23 kb. DNA concentration was measured using the 1x dsDNA High Sensitivity Qubit Assay (Thermo Fisher Scientific), while fragment size distribution was assessed with the Agilent Genomic DNA kit on the Tapestation (Agilent).

##### Library preparation and sequencing

Genomic DNA samples underwent library preparation following the ONT Ligation Sequencing gDNA - Native Barcoding Kit 24 V14 protocol. The process included end-repair, native barcoding, and adapter ligation. Samples were pooled in sets of three, with 800 ng or 1000 ng of input per sample. The final libraries yielded a mean of 56.69 fmol (>35 fmol) per pool.

Sequencing was performed on the PromethION Flow Cell R10 (M version) using the P2 Solo Sequencer (MinION release 24.02.16). The library was divided into three portions and loaded in a tapered manner across three time points. At the first time point, the initial portion of the library was loaded, and sequencing began in whole-genome mode (without adaptive sampling), to monitor QC (N50 within the expected range). After one hour, adaptive sampling was activated. A custom BED file was used to define the target regions (N=326). Each region was extended by 20 kb upstream and downstream, and overlapping regions were merged into non-redundant intervals using bedtools^43^, covering a total of 1.3% of the human genome. At the second time point (20–24 hours after sequencing began, or when fewer than 2000 pores remained active), the flow cell was washed following ONT’s Flow Cell Wash Kit protocol, and the second portion of the library was loaded. At the third time point (40–48 hours), the second portion of the library was retrieved, and merged with the third portion. The flow cell was washed and reloaded with the mixed library. This loading strategy achieved a mean on-target sequencing depth of 25.2x per sample and an average enrichment ratio of 8.5x. The mean on-target N50 post-sequencing was 7,889 bp (**Supplementary Table 3**).

##### Quality control

Details of the QC metrics applied throughout the workflow are provided in **Supplementary Table 9**.

#### 4.2.3 Bioinformatics analysis and reporting

Bioinformatics analysis and variant interpretation utilize a combination of third-party and in-house tools for basecalling, read mapping, variant calling, and pharmacogenomic annotation (Supplementary Figure 11). Additionally, the workflow includes genome-wide imputation from off-target signals. Detailed descriptions of each step are provided below.

##### Basecalling and read mapping

POD5 files were basecalled using Dorado v0.8.1 in either high-accuracy (HAC) or super-accuracy (SUP) mode with the corresponding models (dna_r10.4.1_e8.2_400bps_HAC@v5.0.0 or SUP@v5.0.0). Reads from the initial 1-hour WholeGenome Sequencing (WGS) and subsequent adaptive sampling were filtered for Q-scores >10. Passing reads were demultiplexed using Dorado’s demux function, then combined per sample and mapped to the GRCh38 reference genome using Minimap2 v2.22.

Summary statistics were generated from unmapped BAM files using Dorado’s summary function and processed with custom scripts. Summary metrics for the 1-hour WGS and adaptive sampling stages are presented in **Supplementary Tables 3a** and **3b**, respectively. N50 values were estimated using Cramino v0.14.5^44^ after subsetting reads into on-target and off-target regions. Median coverage depths for these regions were calculated using mosdepth v0.3.6^45^.

### Genotyping of target pharmacogenes

#### Small variants

Small variants (SNPs and indels) were called using Clair3 v1.0.5 with either the *r1041_e82_400bps_hac_v500* or *r1041_e82_400bps_sup_v500* model, as appropriate, and subsequently phased using WhatsHap v2.22.

#### Structural variants

For all pharmacogenes except *CYP2D6*, gene-level copy number was estimated by normalizing the mean on-target depth across each region of interest to the overall mean across all on-target regions. Coverage was calculated using mosdepth v0.3.6 with the--no-per-base option and 100 bp non-overlapping windows.

For hybrid genes (e.g., *CYP2D6/CYP2D7*, *CYP2A6/CYP2A7*), hybrid detection was performed by classifying reads based on distinguishing variants catalogued in PharmVar v6.2.2 or reported by Chen et al. (2021). A hybrid star allele was assigned if at least 3 reads supported the same haplotype.

Due to the complex structural variability of *CYP2D6*, including common duplications, deletions, and gene conversions with *CYP2D7*, its structural analysis is described in detail in a dedicated section.

#### Star-alleles

Based on the Variant Call Format (VCF) calls, star alleles for *CYP2B6*, *CYP2C19*, *CYP2C9*, *CYP3A4*, *CYP3A5, CYP4F2, DPYD, G6PD, NUDT15, SLCO1B1, TPMT* and *UGT1A1* were assigned using PharmCAT^46^ version 2.2.3 with CPIC allele definitions v1.22.2 and star alleles for *NAT2, CYP2C8* and *ABCB1* were assigned using PyPGx version 0.25.0^47^. Definitions for *NAT2* were updated according to PharmVar (accessed: 2024-09-10).

#### HLA typing

HLA calls were made using HLA-LA^48^ version 1.0.3 applying a threshold cutoff of 0.95.

#### CYP2D6 calling

An in-house developed caller was used to resolve diplotype calls in *CYP2D6*. As shown in Supplementary Figure 12, for *CYP2D6*, reads mapping to positions 42,001,498 to 42,255,865 on GRCh38 chromosome 22 were extracted. As a first step, reads corresponding to potential hybrid alleles, if detected, were removed. This was complemented by a downstream analysis step in which all reads were re-examined to identify hybrid structures (see below). The remaining non-hybrid alignments were used as input for Clair3 (v1.0.9), with a reference VCF containing *CYP2D6* variants of interest, and for NanoCaller (v3.6.0). Variant calls from Clair3 and NanoCaller were then merged and phased using WhatsHap (v2.2). The resulting VCF was processed with PharmCAT (v2.2.3) using the “--research-mode cyp2d6,combination” flag, which enables the calling of both full and partial *CYP2D6* diplotypes. Partial diplotypes were only considered in cases where hybrid genes or structural variants were detected, as these are known to reduce variant calling accuracy in the region. When multiple diplotype candidates shared the same PharmCAT match score, the diplotype with the numerically lower star allele designation was reported, along with a warning indicating that multiple interpretations were possible.

Following initial star allele assignment, gene copy number was refined. Duplications were flagged if reads supporting duplication of the *CYP2D6*-like downstream region (REP6) were detected. In cases where a duplication or more than two star alleles—including hybrids— were identified, diplotypes were resolved by evaluating read support for REP6 or REP7 regions. If direct evidence was insufficient, the haplotype background associated with REP6 or REP7 was inferred to determine likely combinations of hybrid and non-hybrid star alleles or duplications. For duplications involving non-identical star alleles, B-allele frequencies of heterozygous SNPs that differentiate between the alleles were used to estimate copy number. For duplications involving identical star alleles, copy number was estimated from normalized read depth. Deletions were called if at least one read spanning a known deletion breakpoint in the *CYP2D6* region was detected, or if the region spanning positions 42,125,000 to 42,135,000 showed a normalized copy number below 1.5.

As described in the “Structural variants” section, regions of reads mapping to CYP2D6 were classified as originating from CYP2D6 or CYP2D7 based on distinguishing variants. Reads showing a switch between the two homologs were classified as hybrids, and assigned star alleles based on switch position, per PharmVar definitions.

Warnings were issued in the following cases: (1) multiple potential diplotypes detected, (2) a partial genotype was reported, (3) copy number estimates from read depth were not supported by structural variant signature reads, or (4) unexpected genotype combinations were observed, suggesting reduced call quality.

### Imputation

Genome-wide genotype imputation was performed in HG005 samples using GLIMPSE2 v2.0.0^30^ and a reference panel comprising 2,504 high-coverage samples from the 1000 Genomes Project. Whole-genome CRAM files from earlier processing steps were utilized as input.

#### 4.2.4 Post-test consultation

The final report includes diplotype calls interpreted into metabolizer phenotypes based on PharmGKB guidelines^49^, along with actionable recommendations from consortia such as CPIC^50^ and DPWG^51^. Results are compiled into a structured PDF for physician review, forming the basis for post-test consultation, where findings are communicated to the patient and integrated into their treatment plan (example report: **Supplementary File 1**).

### 4.2 Validation samples

Genome in a Bottle (GIAB) gDNA samples were obtained from the National Institute of Standards and Technology (NIST). Additional cell line gDNA samples from the GeT-RM program were sourced from the Coriell Institute for Medical Research.

CAP (College of American Pathologists) samples were obtained from previous CAP PGx surveys. These samples were previously analysed in house and submitted to CAP for verification of results, and the verified results were used as a truth set benchmark.

Whole blood samples were obtained from the Molecular Diagnostic Laboratory at Tan Tock Seng Hospital (TTSH), Singapore. Genotype results were based on specific variants detected using microarray technology (Axiom).

For the interference study, a subset of whole blood samples was spiked with >4.5 mg/dL of triglycerides (Merck) to simulate conditions of hyperlipidaemia or elevated blood triglyceride levels, in accordance with the Ministry of Health’s 2016 Hyperlipidaemia Guidelines.

### 4.3 Performance evaluation

Reference data sources used to evaluate calling performance at the variant, diplotype and phenotype levels are described below and summarized in **Supplementary Table 10**.

#### 4.3.1 LOD study - Variant calling performance for SNPs and indels

For small variants (SNPs and indels), hard-filtered Genomic Variant Call Format (gVCF) calls generated by DRAGEN v4.2.7 (available at https://registry.opendata.aws/ilmn-dragen-1kgp) were processed using the *GenotypeGVCFs* module from GATK v4.5.0.0^52^ to extract variants of interest. These were then compared to Clair3 variant calls using custom in-house scripts.

Performance metrics used in our analysis are defined as follows:

- Callability: The proportion of loci with successfully obtained genotype calls relative to the total number of loci evaluated.
- Genotype concordance: The proportion of loci with accurate genotype calls among those that were successfully genotyped.
- Analytical sensitivity: The proportion of variant sites that were correctly detected.
- Analytical specificity: The proportion of non-variant sites that were correctly identified.
- Precision: The proportion of correctly genotyped variants relative to the total number of variants reported.

False negative and false positive calls were manually reviewed. All metrics were calculated on a per-sample basis, and confidence intervals around the mean values were estimated using Student’s *t*-test.

#### 4.3.2 LOD study - Variant calling performance for SVs

Expected SVs were determined using GeT-RM diplotype calls for *CYP2D6* and StellarPGx diplotype calls for *CYP2A6*. In cases where discrepancies between reference and observed calls were noted, results were manually reviewed to confirm the SV status.

Calls for deletions, duplications, and hybrid alleles were classified as:

- True positives if the expected SV was correctly identified by the pipeline for a given sample,
- False negatives if no SV was called when one was expected, and
- False positives if an SV was called when none was expected.

For *CYP2A6*, *46 alleles (3’ UTR conversions) were considered hybrid alleles and evaluated accordingly. Additionally, a *47 allele (a *CYP2A7:CYP2A6* hybrid consisting of exons 1–8 from *CYP2A7*) that was classified by the pipeline *as \**4 (gene deletion) *was* still considered concordant with the expected SV.

Performance metrics were calculated using calls from both genes across all samples and sequencing runs. Confidence intervals were estimated according to the method described by Newcombe and Altman^53^.

#### 4.3.3 Accuracy study - Diplotype concordance in pharmacogenes with complex allele definitions

For HG001, HG01190, NA18861, NA18966, NA19174 and NA19226, GeT-RM calls for *CYP2A6, CYP2B6, CYP2C19, CYP2C8, CYP2C9, CYP2D6, CYP3A4, CYP4F2, DPYD, NAT2, SLCO1B1* and *TPMT* from the consolidated GeT-RM table dated 20240418 ^54,55^ were used as a truth set. For HG00337, HG005, HG01097, HG01572 and HG03259 diplotype calls were compared against PyPGx diplotypes based on dragen v4.2.7 generated CRAM files. For *NAT2,* diplotype definitions were compared taking into account the differences between previous and current allele definitions.

For all samples, *ABCB1, G6PD, NUDT15* and *UGT1A1* calls were compared against PyPGx calls and *HLA-A* and *HLA-B* calls were compared against calls from optitype v1.3.5^56^ based on dragen v4.2.7-generated Compressed Reference-Aligned Mapping (CRAM) files.

Calls were classified as concordant if matching to the truth set. When not matching, calls were considered improved if the TAS-LRS-based call agreed with evidence from manual checks based on IGV plots or based on consensus calls from other programs (see Section 4.3.5 below).

#### 4.3.4 Accuracy study - Phenotype concordance in pharmacogenes with complex allele definitions

Phenotypes were classified as “Concordant to reference” if TAS-LRS-based calls matched the expected phenotype based on the truth sets described above. To account for differences due to an improvement in diplotype calls, calls were classified as “Concordant with updated call” if the reported phenotype matched the expected phenotype based on improved diplotype calls and calls were classified as “indeterminate due to insufficient information” if a phenotype could not be assigned due to a no call for the diplotype or if a call could not be assigned due to the lack of sufficient information on Copy Number Variation (CNV) to distinguish from samples with indeterminate calls due to the underlying diplotype not having a known phenotype.

#### 4.3.5 Accuracy study - Diplotype calling performance across multiple testing platforms

##### Orthogonal short-read data

Star allele and HLA calls from DRAGEN v4.2.7 were obtained from the Illumina DRAGEN 1000 Genomes Project dataset (https://registry.opendata.aws/ilmn-dragen-1kgp; accessed 16 February 2025), except for sample HG005, for which calls were derived using DRAGEN v4.0.3. For PyPGx and StellarPGx comparisons, diplotype calls were generated by processing the corresponding CRAM files from the same dataset using PyPGx (v0.25.0)^47^ and StellarPGx (v1.2.7) ^57^.

##### Orthogonal microarray data

For microarray calls, star alleles and genotypes were called from the Illumina Global Screening Array v3, as previously described^19^.

Calls were considered matching if they were concordant with results from manual inspection of long-read IGV plots or consistent with consensus calls across multiple tools. The percentage of matching calls was calculated as the total number of concordant calls divided by the total number of calls. Callability was defined as the proportion of successfully generated calls relative to the total number of samples evaluated. Both metrics were aggregated across all samples and sequencing runs, and confidence intervals were calculated using the method described by Newcombe and Altman^53^.

#### 4.3.6 Precision study – Reproducibility and repeatability

Assay precision was assessed by evaluating both reproducibility and repeatability of the performance metrics described in Section 4.3.1. Reproducibility was determined by analyzing inter- and intra-run variability across HG001 and HG005 samples sequenced in three independent runs. Repeatability was assessed by reanalyzing the same runs starting from the basecalling step. Variability was quantified by calculating the coefficient of variation (CV) for each performance metric.

#### 4.3.7 Imputation accuracy

To compare imputation accuracy, FASTQ files from five replicates of HG005—generated on the MGI DNBSeq T7 platform —were aligned to the GRCh38 reference genome using DRAGEN-GATK v1.3.0 and subsequently downsampled to 2× coverage to match the average off-target depth of HG005 TAS-LRS samples (N=4). Imputation was performed using GLIMPSE v2.0.0 as described above.

For microarray data, variant calls from five replicates of HG005—generated using the Infinium Global Screening Array v3 (Illumina)—were phased with Eagle v2.4.1 and imputed using Minimac4 v1.0.3 with the 1000 Genomes Project reference panel.

Imputation concordance (R²) was assessed using the GLIMPSE2_concordance script from the GLIMPSE v2.0.0 package, with the following parameters: “--gt-val--min-tar-gp 0.8--ac-bins 1 5 10 20 50 100 200 500 1000 2000 3202”. Genome-wide NIST reference calls were used as the truth set for benchmarking.

## Data Availability

The data supporting the findings of this study are available within the article and its supplementary materials. The underlying sequencing datasets are currently being deposited to the European Nucleotide Archive (ENA) and will be made publicly available upon acceptance of the manuscript.

## Acknowledgments

We thank the clinical operations team at NalaGenetics, including Dr. Latifah Anandari and Kavimozhi Kandasamy, for their support throughout the study. We are also grateful to the laboratory team at Tan Tock Seng Hospital (TTSH), including Sim Wey Cheng, Lim Chia Wei and Grace Toh Li-Xian, for their assistance with sample processing. Special thanks to Dr. Elaine Lo (National University Hospital) for her clinical insights and guidance. We acknowledge the team at Oxford Nanopore Technologies (ONT) - Mavis Tan, Miles Benton, Lin Yang, Simon Dunbar, Lei Tong, Jerald Yam, Nicholas Ong, Vania Wikasa and Germaine Kwok - for their technical support and collaboration.

## Author Contributions

PGHP and YHL contributed equally to this work. PGHP supervised and contributed to developing the software and pipeline, performed data analysis and co-wrote the manuscript. YHL conducted the laboratory work. MIBH contributed to developing the software and pipeline and assisted with data analysis. YM and KNR assisted with data analysis. ANQH supervised the laboratory procedures. AI and LS contributed to funding acquisition and supervised the study. GLL provided clinical samples and supervised the study. MGP conceived the study, secured funding, supervised the project, and co-wrote the manuscript.

## Competing Interests

PG, HL, MI, YM, AQH, KR, AI, LS, and MGP are employees of NalaGenetics. The remaining author declares no competing interests.

## Materials & Correspondence

Correspondence should be addressed to Dr. Mar Gonzalez-Porta.

## Ethics statement

All patient samples were obtained with informed consent in accordance with institutional guidelines. The study protocol was reviewed and approved by the institutional ethics review board at Tan Tock Seng Hospital (Singapore). All methods were carried out in accordance with relevant guidelines and regulations. No additional ethical approval was required for the use of publicly available reference materials.

## Funding

This work was supported by the Enterprise Development Grant (EDG) by Enterprise Singapore.

## Abbreviations

PGx: pharmacogenomics
ADRs: adverse drug reactions
CPIC: Clinical Pharmacogenetics Implementation Consortium
DPWG: Dutch Pharmacogenetics Working Group
PCR: Polymerase Chain Reaction
TAS-LRS: Targeted Adaptive Sampling–Long Read Sequencing
ONT: Oxford Nanopore Technologies
LOD: Limit of Detection
QC: Quality Control
VIP: Very Important Pharmacogene
PharmGKB: Pharmacogenomics Knowledgebase
FDA: Food and Drug Administration
CAP: College of American Pathologists
CLSI: Clinical and Laboratory Standards Institute
CLIA: Clinical Laboratory Improvement Amendments
IVDR: In Vitro Diagnostic Regulation
EQA: External Quality Assessment
SV: structural variant
SNV: single nucleotide variant
CNV: copy number variant
WGS: whole-genome sequencing
HAC/SUP: high-accuracy / super-accuracy
SNPs: single nucleotide polymorphisms
NIST: National Institute of Standards and Technology
1KGP: 1000 Genomes Project
GeT-RM: Genetic Testing Reference Materials
IGV: Integrative Genomics Viewer
DRAGEN: Dynamic Read Analysis for Genetics
GIAB: Genome in a Bottle
MAF: minor allele frequency
GSAv3: Global Screening Array v3
PRS: Polygenic Risk Score
BED: Browser Extensible Data
gDNA: genomic DNA
SPRI: Solid-Phase Reversible Immobilization
dsDNA: double-stranded DNA
VCF: Variant Call Format
gVCF: genomic VCF
CRAM: Compressed Reference-Aligned Mapping
CV(%): coefficient of variation.

---

**Supplementary Figure 1.**
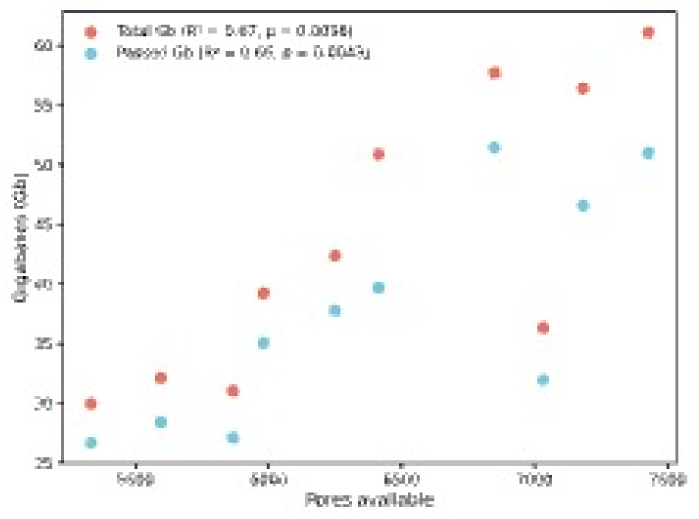
Relationship between flow cell pore availability and sequencing yield. Scatter plot showing sequencing yield (gigabases, Gb) as a function of the number of available pores at the start of each run (n = 10). Each data point represents a sequencing run that included both Whole Genome Sequencing (WGS) and adaptive sampling phases. Total yield (red) and passed yield (blue) are shown. A weak but statistically significant positive correlation (Pearson’s r) was observed for both total (R² = 0.67, p = 0.0036) and passed yield (R² = 0.66, p = 0.0043), indicating that higher pore availability is associated with increased sequencing output.

**Supplementary Figure 2.**
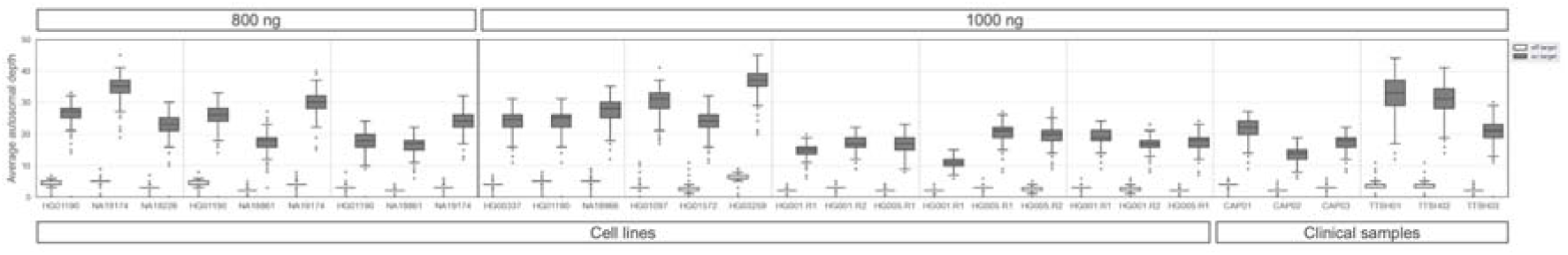
Average autosomal depth of coverage across samples. Boxplots showing the mean depth of coverage for all on- and off-target regions, based on SUP basecalled reads from 3-plex adaptive sampling runs. Samples are grouped by input DNA amount (800 ng and 1000 ng). Boxplots show median, IQR; whiskers extend to 1.5x IQR. Outliers shown as dots.

**Supplementary Figure 3.**
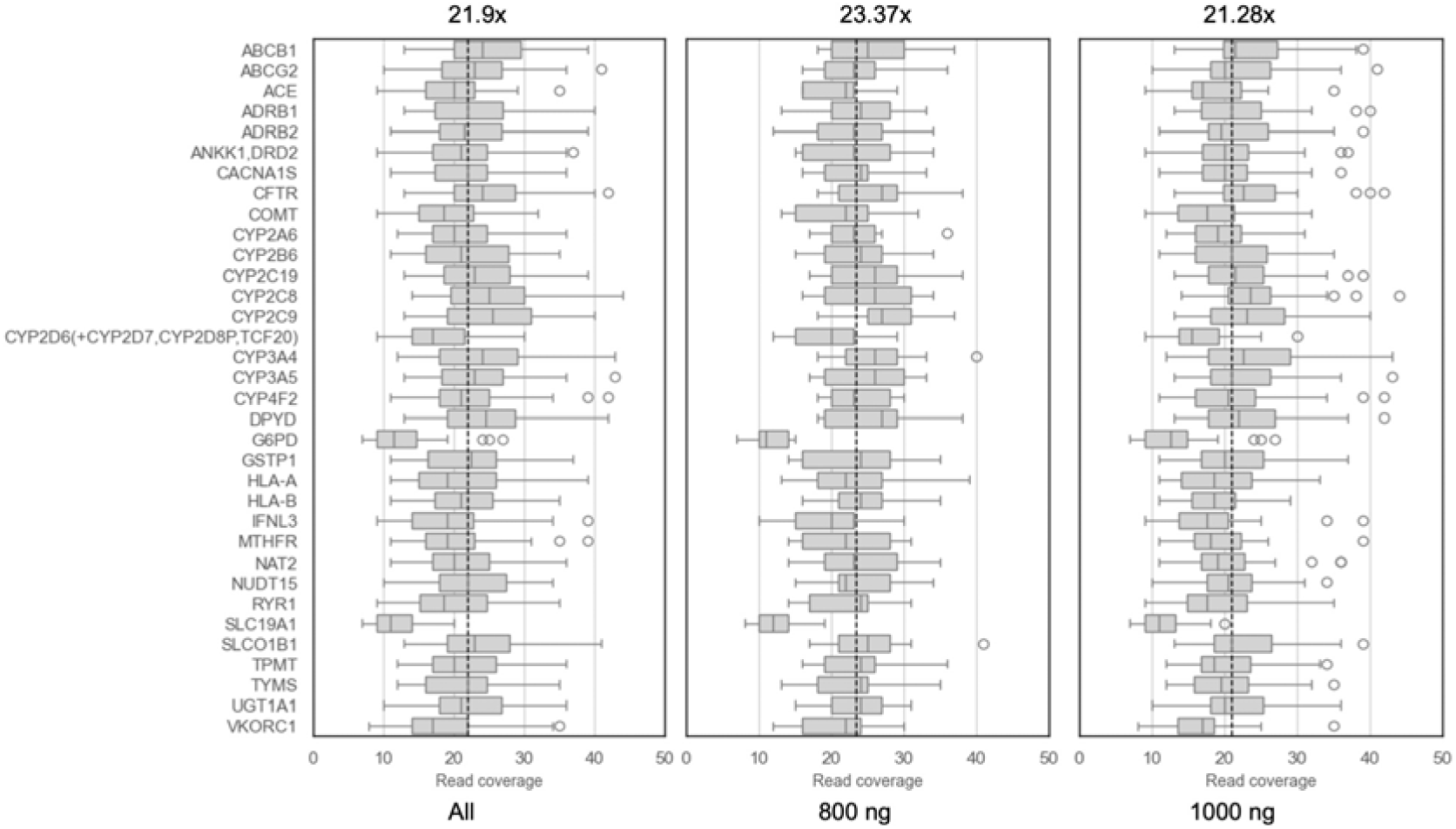
Distribution of read depth in target pharmacogenes. Boxplots representing the distribution of sequencing read depth for 34 pharmacogenes (PGx) targeted in the panel, using a 3-plex adaptive sampling. The dashed vertical line indicates the mean read depth across all genes (21.9x). Most genes demonstrate consistent coverage, while *G6PD* and *SLC19A1* show notably lower coverage across samples. Outliers are depicted as individual points.

**Supplementary Figure 4.**
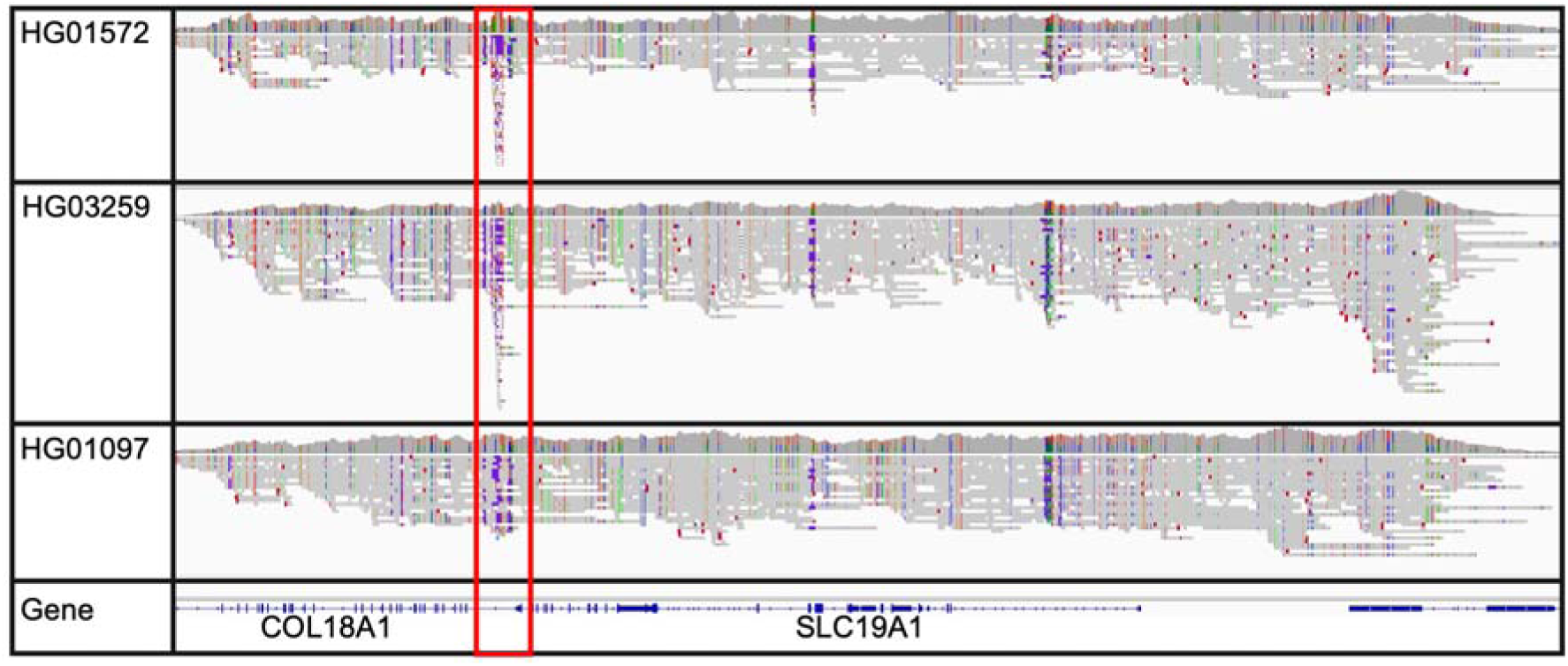
Low-complexity region near *SLC19A1*. IGV genome browser plot showing read alignments for three samples (HG01572, HG03259, HG01097) from the 3-plex run “1000ng_cell_lines_5”, highlighting a low-complexity within *COL18A1*, located adjacent to *SLC19A1* (red box). The gene tracks at the bottom depict the genomic location of *COL18A1* (alpha chain of type XVIII collagen) and *SLC19A1*. The low-complexity region overlaps with the targeted region for adaptive sampling, and reduces the target coverage for *SLC19A1*.

**Supplementary Figure 5.**
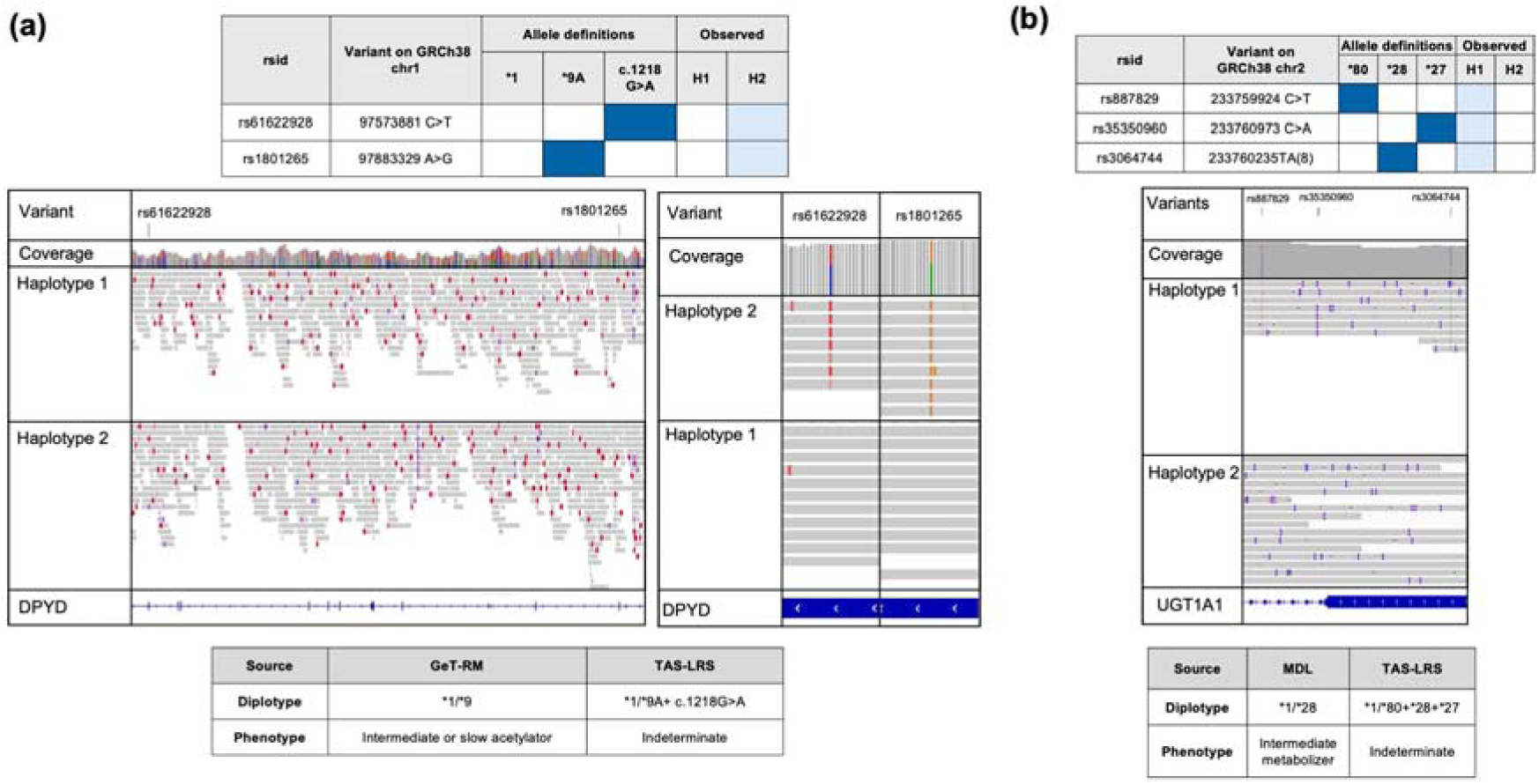
Examples of improved pharmacogene calls enabled by enhanced phasing, novel allele detection, and expanded haplotype coverage. (a) Long-read sequencing enabled phasing of c.85T>C (*9A) and c.1216A>G variants in NA19226. (b) Improved diplotype calling of UGT1A1 in a clinical sample (TTSH_3), where TAS-LRS identified the complex allele *1/*80+*28+*27.

**Supplementary Figure 6.**
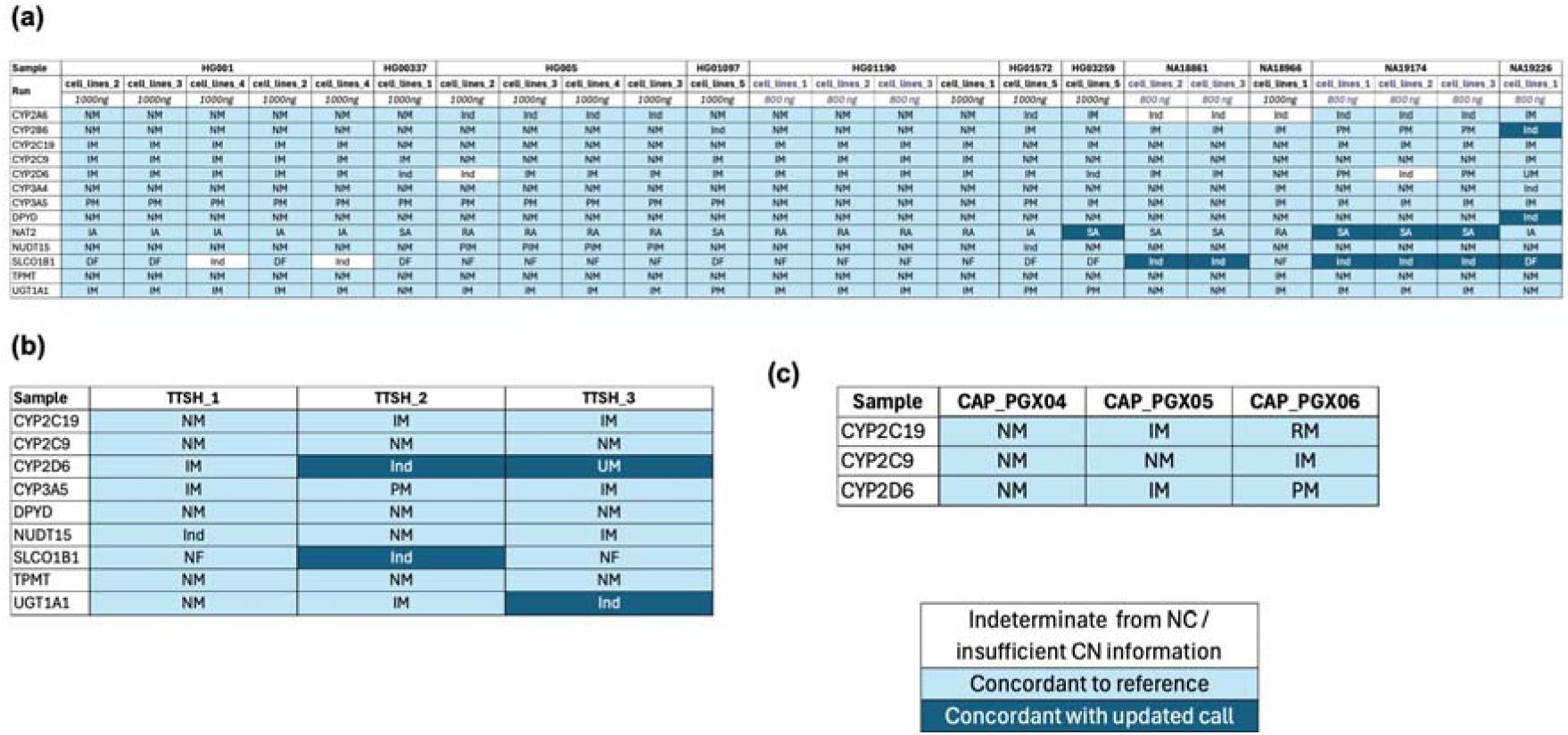
Phenotype concordance in pharmacogenes with complex allele definitions and metabolizer profiles. Results for **(a)** reference cell lines (n=11), **(b)** clinical samples from TTSH (n=3) and **(c)** External Quality Assessment (EQA) samples (n=3) were compared against reference truth sets. Phenotypes were inferred from diplotype calls based on PharmGKB. Phenotypes based on GeT-RM calls were used as the truth set for HG001, HG01190, NA18861, NA18966, NA19174, and NA19226 unless otherwise specified. Phenotype calls for HG00337, HG005, HG01097, HG01572, and HG03259 were based on PyPGx diplotypes from 30x short-read data. For all samples, *NUDT15*, and *UGT1A1* expected phenotypes were based PyPGx diplotypes. Calls matching the expected phenotypes based on the truth set were labelled as concordant, while mismatched calls were considered improved based on IGV read support or consensus across orthogonal methods.

**Supplementary Figure 7.**
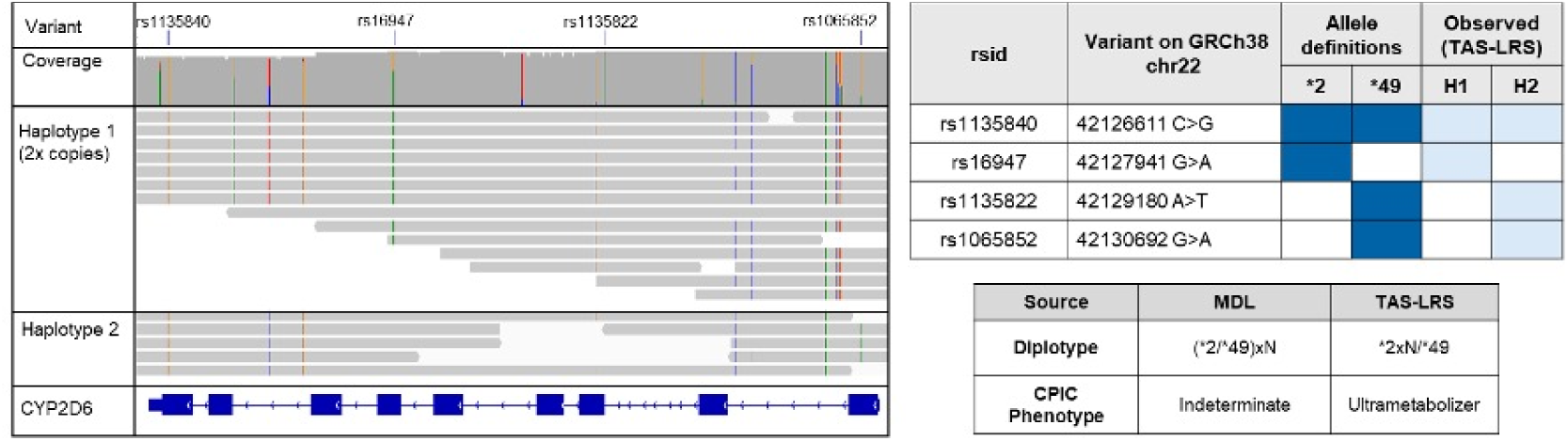
Example of improved *CYP2D6* diplotype call leading to refined phenotype classification. IGV genome browser view and variant table for *CYP2D6* in sample TTSH_3, showing an example where TAS-LRS resolved a previously unphased copy number change identified by microarrays. This allowed for unambiguous diplotype assignment and a change from ‘Indeterminate’ to ‘Ultrarapid metabolizer’ phenotype.

**Supplementary Figure 8.**
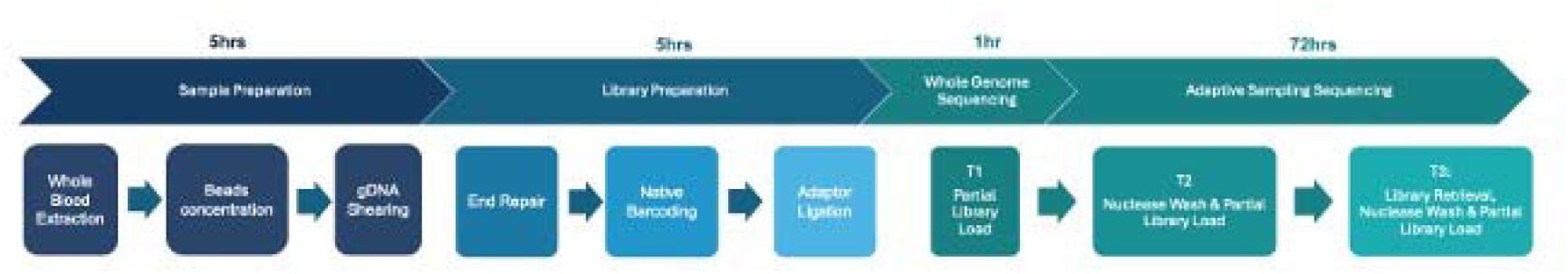
Sample preparation and sequencing workflow for TAS-LRS. Schematic overview of the laboratory protocol used in the TAS-LRS workflow, optimized for three-sample multiplexing. The process includes four main stages: (i) sample preparation (∼5 hours), encompassing whole blood DNA extraction, bead-based concentration, and gDNA shearing; (ii) library preparation (∼5 hours), including end repair, native barcoding, and adapter ligation; (iii) an initial whole genome sequencing (∼1 hour), during which partial libraries are loaded to assess fragment size and flow cell performance; and (iv) adaptive sampling sequencing (∼72 hours), performed in three loading cycles (T1–T3) with nuclease washes and library reloads to maintain active pore utilization.

**Supplementary Figure 9.**
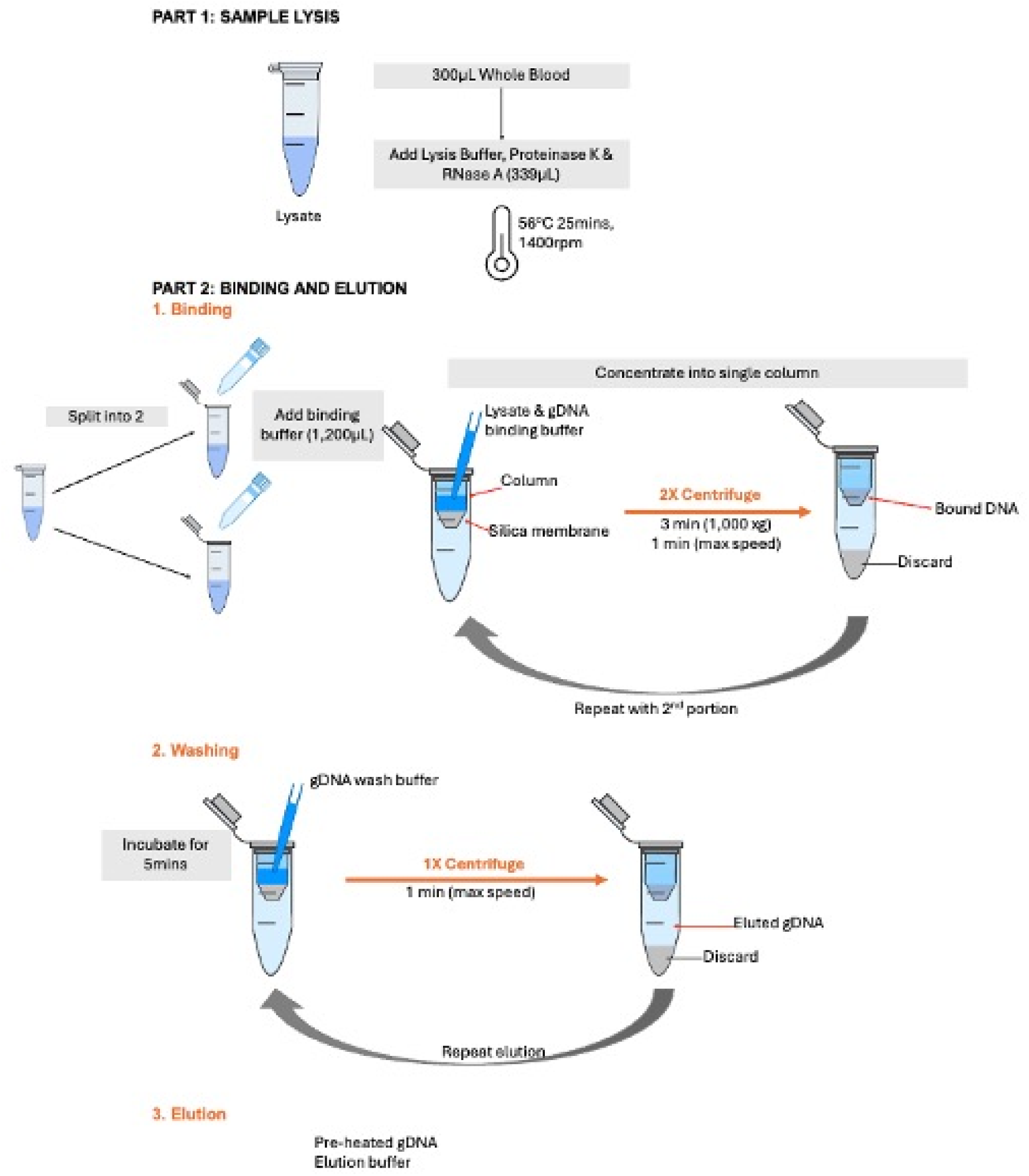
Modified DNA extraction protocol for long-read sequencing. Workflow illustrating the steps modified from the manufacturer protocol to increase recovery of long DNA fragments from blood. Part 1 shows the lysis step, where a scaled-up lysis mastermix is incubated at 56°C for 25 minutes. In Part 2, lysates are split into two portions, with binding buffer and ethanol added prior to column-based DNA binding. DNA is then pooled onto a single spin column, followed by sequential wash steps (Part 3) and a two-step elution using preheated buffer (Part 4) to maximize recovery. Combined with Solid-Phase Reversible Immobilization (SPRI) bead-based concentration, this protocol delivers concentrated gDNA (>100 ng/µL) from 300 µL of blood input.

**Supplementary Figure 10.**
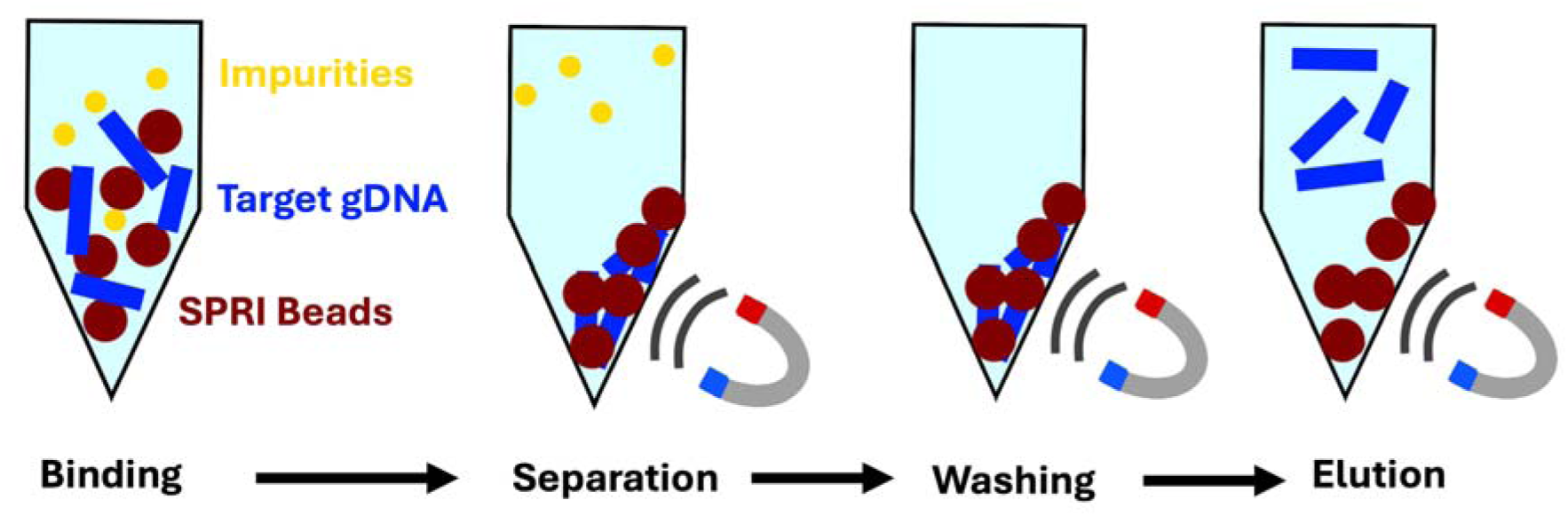
SPRI bead-based concentration of extracted genomic DNA. Diagram of the bead purification workflow used to concentrate genomic DNA following extraction. DNA binds to SPRI beads (dark red) in the presence of polyethylene glycol (PEG) and salt, while impurities (yellow) remain in solution and are discarded during magnetic separation. Subsequent washing steps remove residual contaminants. In the final elution step, purified high-molecular-weight gDNA (blue) is released from the beads into a low-volume buffer, yielding DNA concentrations exceeding 100 ng/µL suitable for long-read sequencing.

**Supplementary Figure 11.**
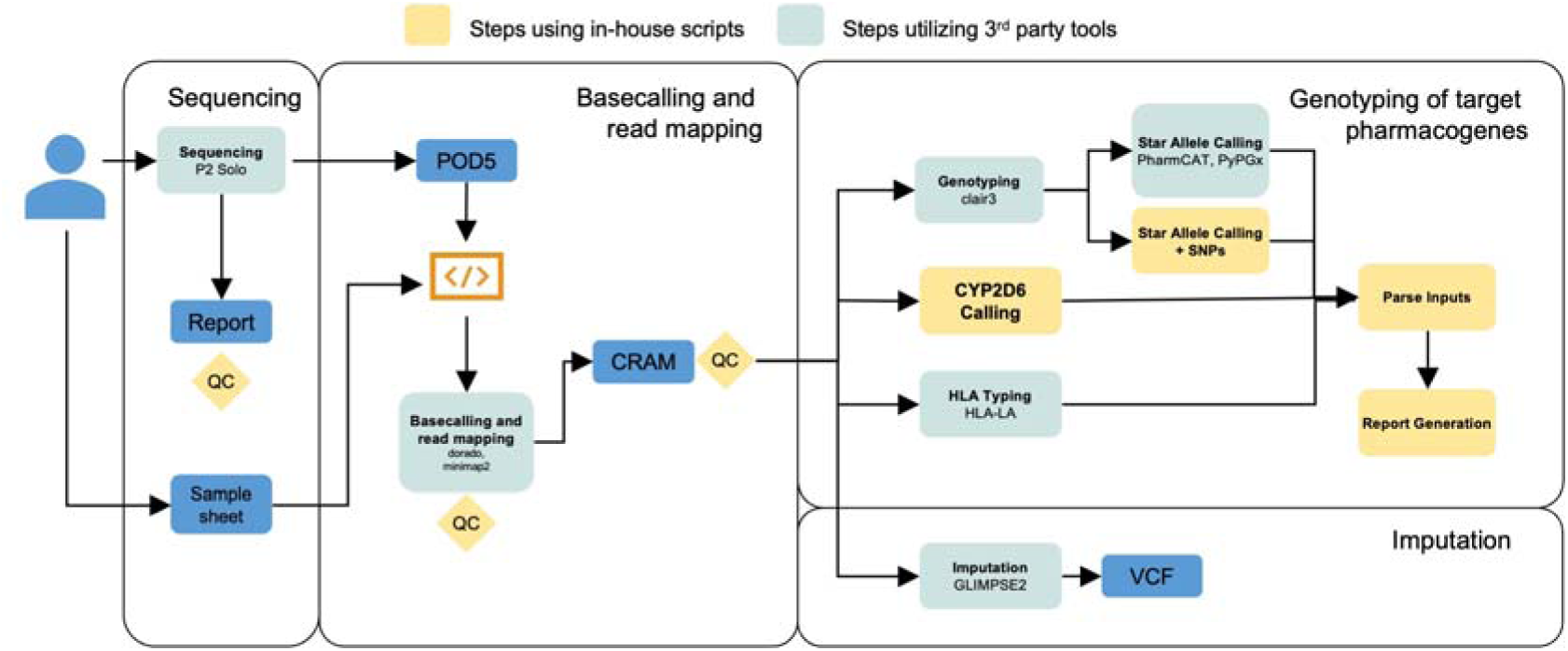
Bioinformatic workflow for pharmacogenomic annotation and genome-wide imputation. Schematic overview of the bioinformatic pipeline, illustrating the steps involved in processing sequencing data for pharmacogenomic analysis. The workflow encompasses sequencing on the ONT P2 Solo platform, basecalling, read mapping, genotyping of target pharmacogenes (including *CYP2D6* and *HLA* typing), and genome-wide imputation using GLIMPSE2. In-house scripts are indicated by orange boxes, while steps utilizing third-party tools are shown in blue. Key file formats - POD5 (raw signal), CRAM (aligned reads), VCF (variant calls) - and quality control (QC) steps are highlighted.

**Supplementary Figure 12.**
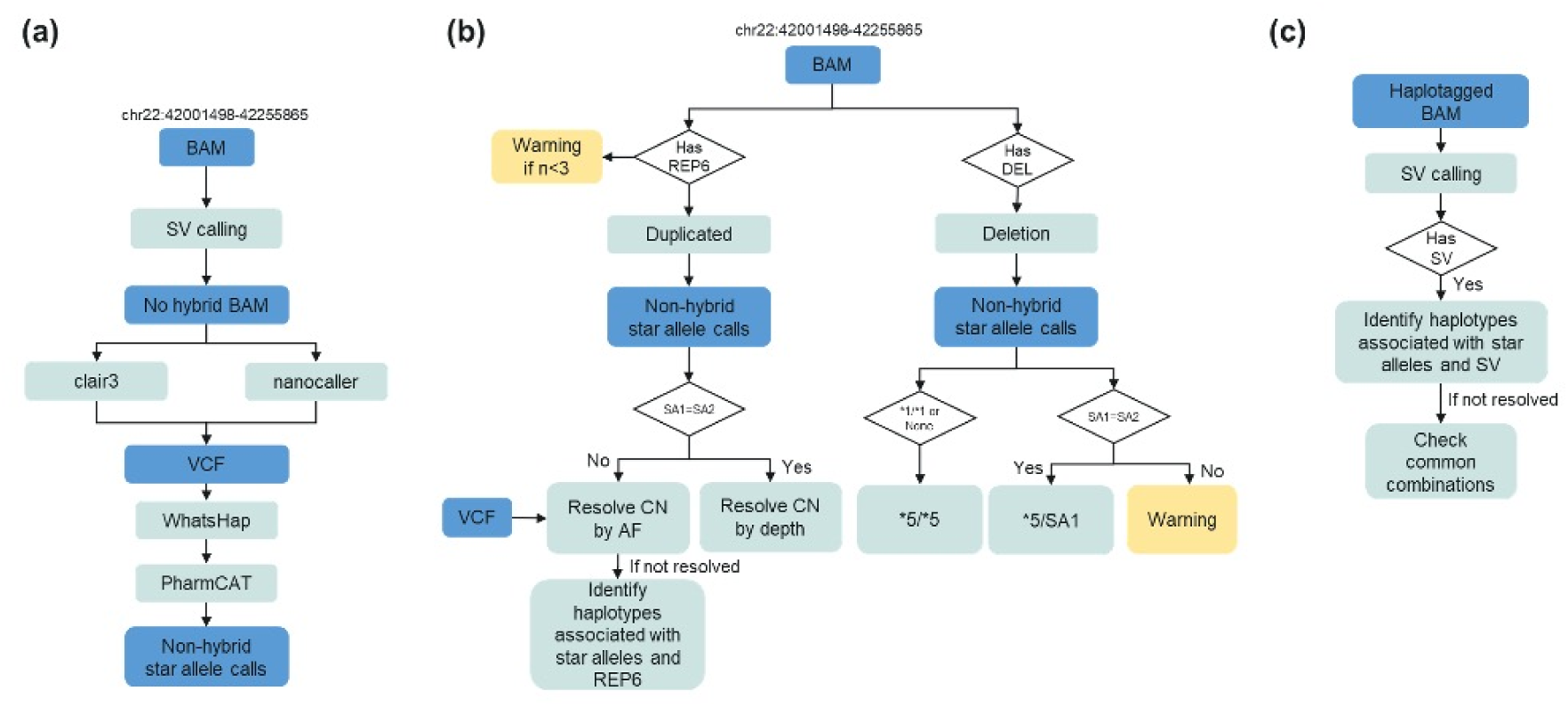
Novel CYP2D6 caller workflow. **(a)** Schematic outlining the initial variant calling process for *CYP2D6*, where non-hybrid reads are used as input for Clair3 and NanoCaller, followed by phasing with WhatsHap and diplotype calling with PharmCAT. **(b)** Heuristic approach for detecting duplications and deletions in *CYP2D6*, including copy number refinement and resolution of complex allele combinations. **(c)** Heuristic for resolving tandem hybrids, involving structural variant (SV) calling and haplotype association to determine hybrid allele composition.

## Table Legends

**Table 1 | Evaluation of small variant genotyping accuracy.** Performance metrics of the limit of detection (LOD) study evaluating small variant calling (SNPs and indels) across two DNA input levels (1000 ng and 800 ng) and two basecalling models (HAC and SUP). A total of 1,142 variants (1,103 SNPs and 39 indels) were analyzed across 31 pharmacogenes, excluding *CYP2D6*, *CYP2A6, HLA-A*, and *HLA-B* due to their high genomic complexity. Sequencing was performed on the Oxford Nanopore Technologies PromethION P2 Solo using a 3-plex sequencing strategy. Metrics shown include callability, concordance, sensitivity, specificity, and precision, each with corresponding 95% confidence intervals (CI).

**Table 2 | Evaluation of structural variant detection performance.** Evaluation of structural variant (SV) detection performance of deletion, duplication, and hybrid-calling in *CYP2D6* and *CYP2A6* genes across all samples by DNA input amount (800 ng and 1000 ng) and by basecalling model (HAC and SUP). Metrics are reported with 95% confidence intervals based on Newcombe and Altman (2000). TN: true negatives, TP: true positives, FP: false positives, FN: false negatives, NC: no-calls.

**Supplementary Table 1 | Pharmacogenes included in the TAS-LRS panel.** This table lists all pharmacogenes included in the TAS-LRS panel. Thirty-four genes were selected based on their classification as Very Important Pharmacogenes (VIP) in PharmGKB. HLA-A was additionally included since it is routinely tested in our laboratory.

**Supplementary Table 2 | Samples used for TAS-LRS workflow validation.** Overview of samples used for validation of the TAS-LRS workflow. Samples were selected to cover a range of structural variants and diplotype configurations relevant to pharmacogenes, particularly CYP2D6. Reference samples from GIAB, 1KGP, and GeT-RM (n=11) were used for benchmarking small variants (SNVs and indels), large structural variants (SVs), and diplotype calls. Additional samples from CAP EQA (n=3) and clinical cases (n=3) were included to assess concordance with orthogonal testing methods.

**Supplementary Table 3 | Quality control and sequencing metrics for study samples in 3-plex runs.** Summaries of sequencing performance and QC metrics for cell line and clinical research samples processed with 800 ng and 1000 ng input amounts. Metrics are reported separately for (a) the 72-hour adaptive sampling phase and (b) the 1-hour whole genome sequencing phase.

**Supplementary Table 4 | Structural variant detection in samples with >25x depth.** Evaluation of structural variant (SV) detection performance of deletion, duplication, and hybrid-calling in *CYP2D6* and *CYP2A6* genes across samples with a greater than 25x on-target depth by DNA input amount (800 ng and 1000 ng) and by basecalling model (HAC and SUP). Metrics are reported with 95% confidence intervals based on Newcombe and Altman (2000). TN: true negatives, TP: true positives, FP: false positives, FN: false negatives, NC: no-calls.

**Supplementary Table 5 | Cross-platform comparison of pharmacogene callability and accuracy.** Percentage of matching diplotype calls using manual or consensus calls as truth sets and callability for 20 pharmacogenes across five platforms: microarray (Illumina GSA v3), short-read whole genome sequencing (WGS) using Dragen, StellarPGx and PyPGx and long-read sequencing (TAS-LRS). Results are presented as percentages with 95% confidence intervals. Sample sizes (n) are indicated for each platform. NA indicates data not available. TAS-LRS generally demonstrated higher concordance and callability across genes (e.g., *CYP2D6*, *UGT1A1*, and *DPYD*), highlighting the advantages of long-read approaches in pharmacogenomic profiling.

**Supplementary Table 6 | Reproducibility of variant calling. (a)** Inter-run and **(b)** intra-run coefficients of variation (CV) of HG001 and HG005 replicates across three independent sequencing runs for callability, genotype concordance, sensitivity, specificity, and precision of 1,142 variants (1,103 SNPs and 39 indels) across 31 pharmacogenes, excluding *CYP2D6*, *CYP2A6, HLA-A*, and *HLA-B*.

**Supplementary Table 7 | Evaluation of DNA extraction consistency and concentration fold change using spiked samples.** This table summarizes DNA quantification metrics for three blood samples (TTSH_4, TTSH_5, TTSH_6) spiked with synthetic DNA at 4,500 ng/µL. For each sample, Nanodrop and Qubit measurements were recorded before and after SPRI bead concentration. Fold change after bead purification and between spiked and non-spiked conditions was calculated. One replicate (TTSH_4) was excluded from analysis due to improper mixing prior to extraction. Mean, standard deviation (SD), and coefficient of variation (CV) are provided across replicates to assess consistency.

**Supplementary Table 8 | Cross-contamination estimates from VerifyBamID2.** Values for contamination correspond to FREEMIX values representing the estimated proportion of foreign DNA present in each sample.

**Supplementary Table 9 | Quality control metrics and acceptance criteria for library preparation and sequencing.** Summary of key quality control (QC) metrics and corresponding acceptance criteria applied at each stage of the workflow, encompassing sample preparation, library construction, sequencing, and post-sequencing analysis. The metrics include assessments of genomic DNA (gDNA) quality and yield, library size and concentration, sequencing performance, and potential contamination. Acceptance criteria are provided to ensure the generation of high-quality sequencing data suitable for downstream analyses. TAS-LRS: Targeted Amplicon Sequencing - Long Read Sequencing; WGS: Whole Genome Sequencing; NBD: Native Barcoding.

**Supplementary Table 10 | Reference datasets for benchmarking variant and diplotype calls.** External truth datasets used to evaluate the accuracy and callability of TAS-LRS across pharmacogenes and HLA loci. For each sample, the table indicates the corresponding validation experiment, the data type (e.g., gVCF, CRAM), and the associated data source path.

## Notes

### Summary of Updates

Added new version of the manuscript, with an additional Supp Figure.

