## Supplementary File 1 for "Targeted adaptive sampling enables clinical pharmacogenomics testing and genome-wide genotyping"

### Clomipramine

MODERATE RECOMMENDATION

| GENE | GENOTYPE | PHENOTYPE |
|---|---|---|
| CYP2C19 | *1/*1 | CYP2C19 Normal Metabolizer |
| CYP2D6 | *1/*4 | CYP2D6 Intermediate Metabolizer |

#### Recommendation

**Consider a 25% reduction of recommended starting dose.**

CYP2D6 IM: Reduced metabolism of tricyclic antidepressants (TCAs) to less active compounds. Higher plasma concentrations of active drug may increase probability of side effects. CYP2C19 NM: Normal metabolism of tertiary amines.

#### Recommendations based on specific guidelines

| | |
|---|---|
| CPIC | Consider a 25% reduction of recommended starting dose. |

#### Caveat

The application of genotype-based dosing is most appropriate when initiating therapy with a tricyclic. Obtaining a pharmacogenetic test after months of drug therapy may be less helpful in some instances, as the drug dose may have already been adjusted based on plasma concentrations, response, or side effects. Similar to all diagnostic tests, genetic tests are one of several pieces of clinical information that should be considered before initiating drug therapy.

#### Source

Publications related to local relevance for this drug-gene pair have not been published yet.

**Decrease Starting Dose**
Patient has altered metabolism rate or enhanced activation rate for drug indicated. Decreasing starting dose has shown to help prevent patient from experiencing adverse drug reaction.

**Initiation**
This drug-gene interaction recommendation is useful for initiation of this drug in naive patients. For patients who are already on stable dose or have used this drug before, effective drug monitoring and clinical judgement is more important.

This report was validated and generated automatically. No signature is required. Recommendations given in this report are made using a lab developed test which should not supersede clinical judgement or medical expertise.
